# Early and rapid identification of COVID-19 patients with neutralizing type I-interferon auto-antibodies by an easily implementable algorithm

**DOI:** 10.1101/2021.11.12.21266249

**Authors:** Bengisu Akbil, Tim Meyer, Paula Stubbemann, Charlotte Thibeault, Olga Staudacher, Daniela Niemeyer, Jenny Jansen, Barbara Mühlemann, Jan Doehn, Christoph Tabeling, Christian Nusshag, Cédric Hirzel, David Sökler Sanchez, Alexandra Nieters, Achim Lother, Daniel Duerschmied, Nils Schallner, Jan Nikolaus Lieberum, Dietrich August, Siegbert Rieg, Valeria Falcone, Hartmut Hengel, Uwe Kölsch, Nadine Unterwalder, Ralf-Harto Hübner, Terry C. Jones, Norbert Suttorp, Christian Drosten, Klaus Warnatz, Thibaud Spinetti, Joerg C. Schefold, Thomas Dörner, Leif Sander, Victor M. Corman, Uta Merle, Pa-COVID study Group, Florian Kurth, Horst von Bernuth, Christian Meisel, Christine Goffinet

## Abstract

**Purpose:** Six-19% of critically ill COVID-19 patients display circulating auto-antibodies against type I interferons (IFN-AABs). Here, we establish a clinically applicable strategy for early identification of IFN-AAB-positive patients for potential subsequent clinical interventions.

**Methods:** We analysed sera of 430 COVID-19 patients with severe and critical disease from four hospitals for presence of IFN-AABs by ELISA. Binding specificity and neutralizing activity were evaluated via competition assay and virus-infection-based neutralization assay. We defined clinical parameters associated with IFN-AAB positivity. In a subgroup of critically ill patients, we analyzed effects of therapeutic plasma exchange (TPE) on the levels of IFN-AABs, SARS-CoV-2 antibodies and clinical outcome.

**Results:** The prevalence of neutralizing AABs to IFN-α and IFN-ω in COVID-19 patients was 4.2% (18/430), while being undetectable in an uninfected control cohort. Neutralizing IFN-AABs were detectable exclusively in critically affected, predominantly male (83%) patients (7.6% IFN-α and 4.6% IFN-ω in 207 patients with critical COVID-19). IFN-AABs were present early post-symptom onset and at the peak of disease. Fever and oxygen requirement at hospital admission co-presented with neutralizing IFN-AAB positivity. IFN-AABs were associated with higher mortality (92.3% versus 19.1 % in patients without IFN-AABs). TPE reduced levels of IFN-AABs in three of five patients and may increase survival of IFN-AAB-positive patients compared to those not undergoing TPE.

**Conclusion:** IFN-AABs may serve as early biomarker for development of severe COVID-19. We propose to implement routine screening of hospitalized COVID-19 patients according to our algorithm for rapid identification of patients with IFN-AABs who most likely benefit from specific therapies.

## INTRODUCTION

Since its first detection in Wuhan, China, in 2019, Severe-acute-respiratory syndrome Coronavirus 2 (SARS-CoV-2) has placed an unprecedented burden on health care systems worldwide. The clinical spectrum of the associated disease, COVID-19, ranges from asymptomatic infection to severe disease with hypoxemia, acute respiratory distress syndrome (ARDS), multiorgan failure, and death (Pfortmueller et al. 2021). Approximately 35% of patients remain asymptomatic, 55% develop upper respiratory tract infections, whereas 15% develop severe pneumonia (defined as SpO2 < 90% at room air) and 5% critical pneumonia (defined as Acute Respiratory Distress Syndrome (ARDS), requiring mechanical ventilation or extra-corporeal membrane (ECMO) (Sah et al. 2021).

Scores containing clinical and laboratory parameters support risk stratification and resource allocation in clinical practice worldwide (Knight et al. 2020; “Information on COVID- 19 Treatment, Prevention and Research” n.d.). Demographic and clinical risk factors for a severe disease course include advanced age, male sex, and pre-existing comorbidities (Williamson et al. 2020). Moreover, genetic polymorphisms are associated with progression to severe disease (COVID-19 Host Genetics Initiative 2021). Cell-intrinsic innate viral sensors and antiviral cytokines, including type I and type III interferons (IFNs), orchestrate the control of SARS-CoV-2 infection (Kim and Shin 2021). Inherited mutations of genes involved in IFN induction and signalling and circulating auto-antibodies (AABs) that neutralize type I IFNs have been found to predispose infected individuals to severe COVID- 19 (Bastard et al. 2020; Zhang et al. 2020), presumably by contributing to an ineffective immune response with delayed or abolished type I IFN signaling. Neutralizing type I IFN- AABs are present in 6-17% of hospitalized COVID-19 patients with severe pneumonia (Bastard et al. 2020; Koning et al. 2021; Troya et al. 2021) and 11-19% in critically ill COVID-19 patients (Bastard et al. 2020; van der Wijst et al. 2021; Goncalves et al. 2021), greatly exceeding estimated prevalences of around 0.33% (Bastard et al. 2020) in uninfected individuals. Intriguingly, while neutralizing IFN-AABs in patients with autoimmune polyendocrine syndrome type 1 (APS-1) can associate with a severe course of SARS-CoV-2 infection (Bastard et al. 2021; Beccuti et al. 2020; Carpino et al. 2021; Ferré et al. 2021; Lemarquis et al. 2021), their mere presence does not inevitably lead to severe disease (Meisel et al. 2021). A recent global multi-cohort study reports prevalence of neutralizing IFN-AABs in 4% of uninfected individuals over 70 years of age, suggesting that IFN-AABs may pre-exist in some individuals that develop a critical course of COVID-19 (Bastard et al. 2021). Thus, we reasoned that IFN-AABs may serve as biomarkers that could, in conjunction with other clinical parameters, help to predict risk for developing severe COVID-19 and to stratify patients for specific therapies.

Specific therapies may comprise the administration of recombinant IFN-β or therapeutic plasma exchange (TPE). However, the clinical benefit of TPE and other approaches remains to be defined and requires studies involving large numbers of patients. With IFN-AABs present in up to 18% of deceased COVID-19 patients (Bastard et al. 2021) and given the limited therapeutic options for severely affected COVID-19 patients, testing specific therapeutic approaches is of high urgency, yet clinically implementable strategies for rapid and early identification of IFN-AAB-positive patients upfront are missing.

## METHODS

### Study cohorts and data collection

Patients were recruited and data and sample collection was performed within one of four prospective observational studies conducted at Charité - Universitätsmedizin Berlin, Germany (Cohort A, (Kurth et al. 2020)), Inselspital Universitätsspital Bern, Switzerland (Cohort B), Universitätsklinikum Freiburg, Germany (Cohort C) and Universitätsklinikum Heidelberg, Germany (Cohort D). For this analysis, all patients with a maximum WHO score of 3-8 were included from Cohorts A-C (henceforth summarized as cross-sectional cohorts, CSC). For cohort D (therapeutic plasma exchange cohort, TPEC) only patients who underwent therapeutic plasma exchange for treatment of COVID-19-associated hyperinflammatory syndrome at the Department of Internal Medicine IV of Heidelberg University Hospital, Germany (Nusshag et al. 2021) and supplementary methods) were retrospectively selected. All TPE procedures were performed in accordance with the German Medical Devices Act (“Medizinproduktegesetz”). Healthy controls were recruited from a study on SARS-CoV-2 exposition in health care workers (HC cohort). Samples from APS-1 patients were obtained from a published study (Meisel et al. 2021) and published values are shown here for reference. All studies were conducted according to the Declaration of Helsinki and Good Clinical Practice principles (ICH 1996).

### Detection of IFN-AABs by reverse ELISA

IFN-AABs were detected using an electrochemiluminescence immunoassay (ECLIA)- platform (MSD, Rockville, U.S.), as described recently (Meisel et al. 2021). Briefly, MSD GOLD 96-well small spot streptavidin SECTOR Plates (MSD) were washed with wash buffer (MSD), and blocked with 150 μl blocking buffer (Thermo Fisher, Waltham, U.S.) per well at 4°C overnight. All further incubations were performed for 60 minutes at room temperature.

After blocking, plates were incubated with IFN-α2 (Merck Sharp & Dohme, Kenilworth, USA) or IFN-ω (Peprotech, Rocky Hill, USA) linked to biotin (Thermo Scientific, Waltham, USA).

Next, plates were incubated with patients’ sera following dilution at 1:100 in blocking buffer. Cytokine AABs were detected using a monoclonal mouse antibody to human IgG (D20JL-6, MSD). After incubation and washing, 150 μl of read buffer (ReadBufferT (4x), MSD) were added, incubated for ten minutes at room temperature, and plates were analyzed using the MESO QuickPlex SQ 120 analyzer (MSD). Data are shown as light signal counts (LSC).

### Competition assays

All sera whose IFN-α2-AABs and/or IFN-ω-AABs levels exceeded the 97.5th percentile of AAB levels of the analysed HCW sera and 27 samples that scored close to, but below this cut-off were assessed by competition assay using unbiotinylated IFN-α2 or IFN-ω. The sera of Interest were diluted 1:100 with blocking buffer and Incubated overnight at 4^0^C with 2.5mg∕ml, 0.025mg∕ml, and 0.00025mg∕ml u∩blotl∩ylated IFN-α2 or IFN-ω. After Incubation, reverse ELISA was performed, as described above. IFN-AABs In a given serum scored specific when prel∩cubatlo∩ with the highest concentration of IFN-α2 or IFN-ω resulted In an at least four-fold reduction of LSC In comparison to analysis of the Identical serum without IFN-α2 or IFN-ω pre-incubation.

### Virus infection-based neutralization assays

Calu-3 cells were pre-incubated with 1% human serum in the presence or absence of 200­400 IU/ml IFN-α2a (Roferon®-A, Roche) or 20-50 ng/ml IFN-ω (PeproTech). After 24 hours, IFN and serum were removed and cells were infected with SARS-CoV-2 at MOI 0.01. Virus inoculum was removed after one hour, cells were washed with PBS and 100 μl medium was added per well. 24 hours post-infection, cell culture supernatant was collected for viral RNA quantification by RT-PCR and infectious titer determination by plaque assay.

### Cytokine and chemokine measurements

Cytokines and chemokines from a subset of patients from cohort A were analyzed using Quanterix’ single molecule array technology or multiplex ECLIA.

## RESULTS

We analyzed 430 serum samples collected within four independent observational clinical studies on COVID-19 for IFN-AAB positivity (Table 1). Median age of patients in the CSC was 61 years (IQR 52-71) and 72.2% (291/403) were male. Median Charlson Comorbidity Index (CCI) was 3 (IQR 1-4). 27 patients with critical disease course (median max. WHO score 7 (IQR 7-8)) who underwent TPE as compassionate use were selected retrospectively from center D (TPEC). Median age of patients in the TPEC was 65 years (IQR 56-72), 74.1 % (20/27) were male and median CCI was 4 (IQR 3-5). All patients from the TPEC required invasive mechanical ventilation (IMV), 77.8% (21/27) hemodialysis, one patient was treated with extracorporeal membrane oxygenation (ECMO), and 13 out of 27 (48.2%) patients died despite maximum care. 667 serum samples from a healthy cohort (HC) consisting of health­care workers (Table S1) were screened for the presence of neutralizing IFN-AABs to set the cut-off for IFN-AAB positivity.

**Table 1:**
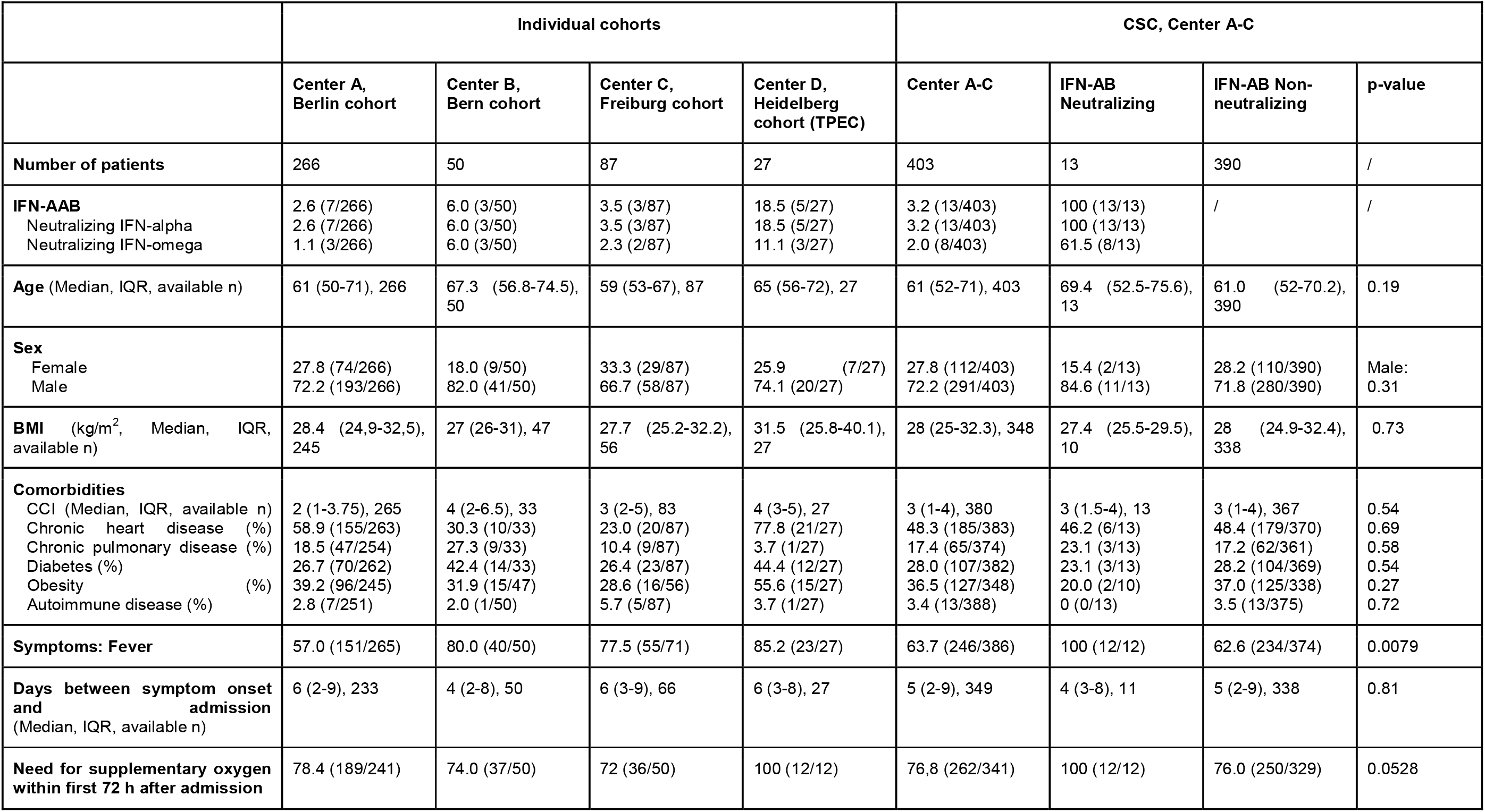
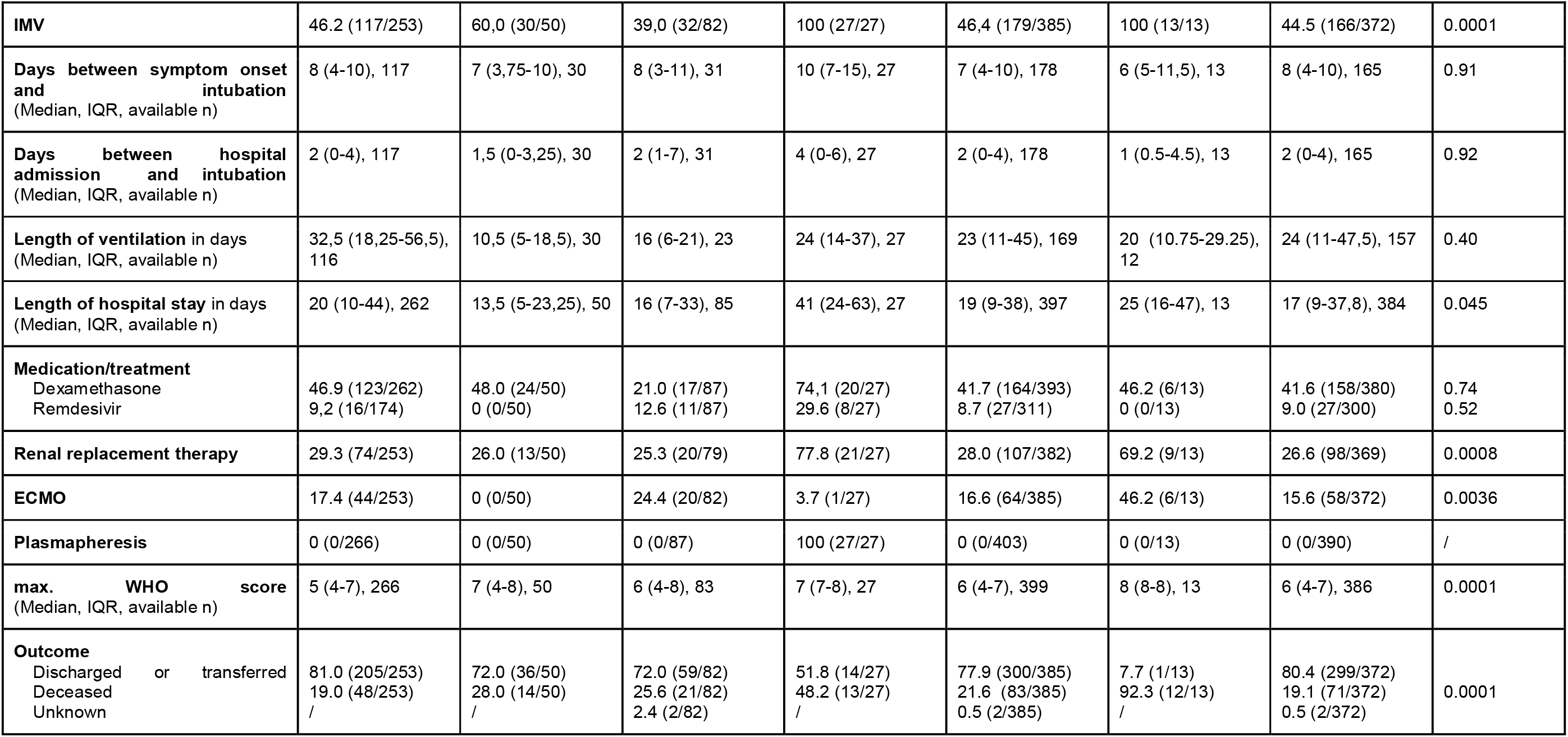
Baseline patient characteristics Data are shown in % (N/n) unless otherwise indicated. IMV - Invasive mechanical ventilation, IQR - interquartile range, CCI Charlson’s comorbidity index Patients with DNI/DNR were excluded for IMV, RRT, ECMO, and Outcome

Prevalence of AABs against IFN-α2 and IFN-ω in patients with COVID-19 We first aimed to establish a sensitive screening assay for type I IFN-AABs. To this end, we first screened samples of our HC for prevalence of AABs against IFN-α and/or IFN-ω by ELISA. Samples were considered positive when the respective LSC value exceeded the 97.5th percentile of AAB levels of the analyzed sera from the HC (cut-off for IFN-α = 1980 LSC, IFN-ω = 1961 LSC). We then screened sera obtained at the peak of the disease from patients of cohorts A-C (CSC) and cohort D (TPEC). The proportion of ELISA-positive patients in the CSC was 5.0% (20/403) for IFN-□ AABs and 4.2% (17/403) for IFN-ω AABs. It was significantly higher in cohort D (TPEC) (IFN-α AABs 18.5%, 5/27, p = 0.0035 and IFN- ω AABs 14.8%, 4/27, p = 0.0132), as expected (Fig. 1a, Fig. S1). Some sera displayed values approaching or equaling those detected In sera from patients with APS-1, a genetic disease Involving the generation of high titer neutralizing type I IFN-AABs (Fig. 1a, (Meisel et al. 2021)).

**Figure 1.**
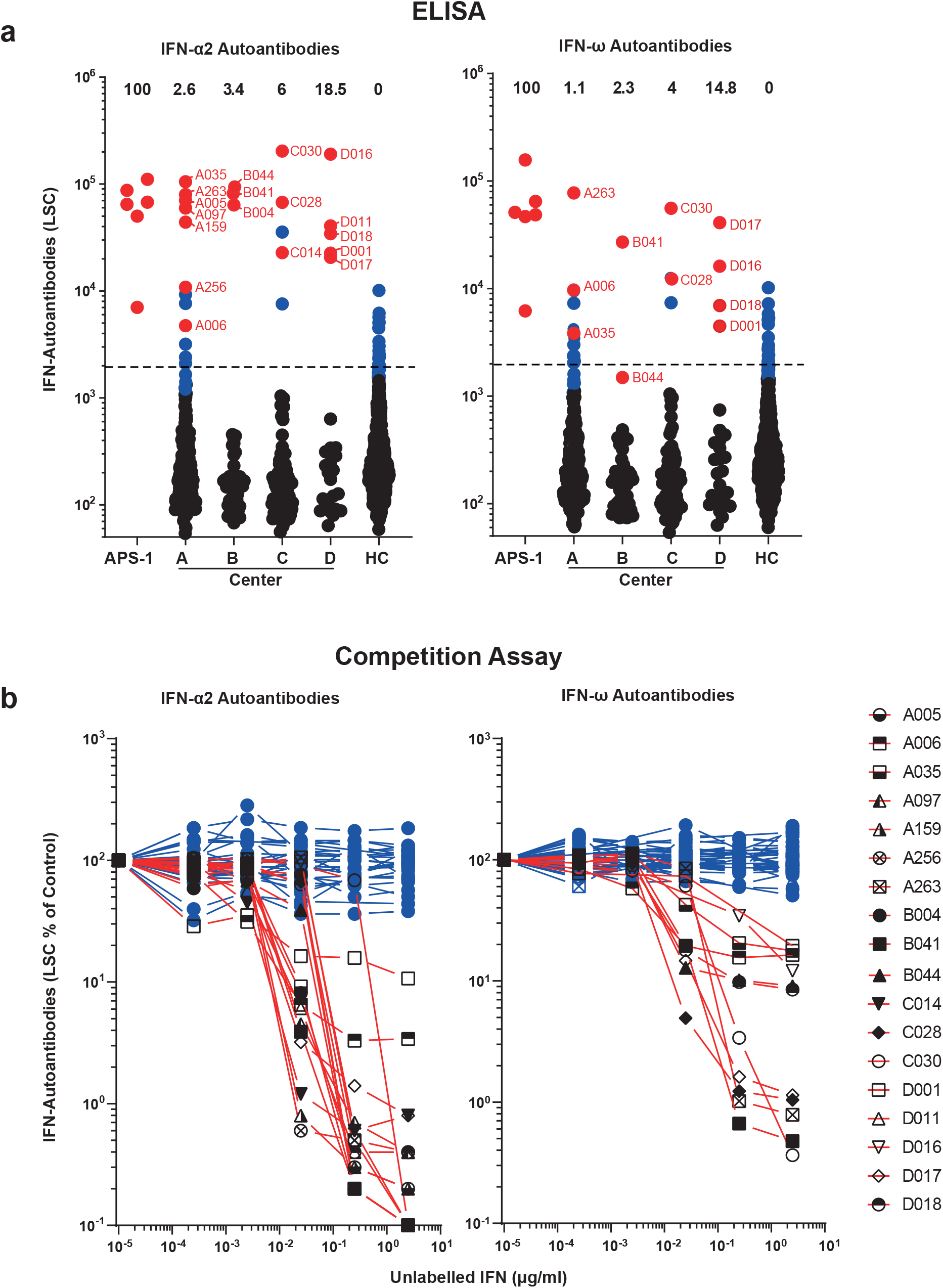
Prevalence of AABs against IFN-α2 and IFN-ω in patients with COVID-19. (a) ECLIA-based assay for detection of IgG AABs against IFN-α2 and IFN-ω in sera from hospitalized patients with COVID-19 from four different university hospital cohorts (Center A, n=266; Center B, n=50; Center C, n=87; Center D, n=27), in patients with APS-1 (n=6), and healthy health care workers (HC) without documented SARS-CoV-2 infection (n=667). Dotted lines indicate the 97.5th percentile of the ECLIA assay LSC in sera from the HC cohort. Dots indicate samples containing AABs scoring specific (red) or unspecific (blue) for IFN-α2 and IFN-ω binding in the competition assay (see b), respectively. The prevalence of sera with specifically binding type I IFN-AABs in each cohort is given in percent. (b) Specificity of the ECLIA assay signal for IFN-α2- and IFN-ω-AABs was tested in an competition assay by preincubation of sera with increasing concentrations of unlabeled IFN- α2 and IFN-ω protein (0 - 2.5 μgλml) before analysis. Samples showing a decrease in assay signal by at least 75% in the presence of the highest competitor concentration were defined as specific for type I IFN antibody reactivity and are indicated with red lines (IFN-α2 n=18, IFN-ω n=11). Samples showing no decrease in the presence of excess unlabeled type I IFN protein were regarded as unspecific for type I IFN antibody reactivity and are indicated with blue lines (IFN-α2 n=62, IFN-ω n=39).

Nonspecific binding is a common phenomenon in immunoassays for the detection of AABs, and high levels of inflammatory parameters such as CRP correlate with non-specific binding of (auto-)antibodies (Güven et al. 2014). Therefore, we probed the specificity of all samples exceeding the 97.5th percentile of the IFN-AAB ELISA and 27 samples that scored right below the cut-off in a competition assay (Fig. 1b). Based on the assumption that sera scoring below the cut-off did not contain IFN-AABs, we established a prevalence of specific IFN-□-AAB of 3.2% (13/403) and of specific IFN-ω-AAB of 1.7% (7/403) in the CSC (Fig. 1a- b). Cohort D (TPEC) showed 18.5% of sera specifically binding IFN- (5/27) and 14.8% for IFN-ω (4/27) or both (14.8%, 4/27) (Fig. 1a-b). As expected, all tested samples scoring below the 97.5th percentile in the ELISA scored negative in the specificity assay with the exception of one patient (B044) who scored slightly below the cut-off in the IFN-ω-AAB assay. Importantly, none of the tested sera from the HC displayed antibodies that specifically bound IFN-□ or IFN-□.

### IFN-AABs neutralize exogenous IFN in a virus infection-based assay

We next analyzed whether the presence of detectable and specifically IFN-binding AABs corresponded to a functional neutralization of IFN during infection. To this end, we applied a previously established assay of IFN-based inhibition of SARS-CoV-2 infection of the immortalized lung cell line Calu-3 (Meisel et al. 2021), which we consider the gold standard for analysis of IFN neutralization. We tested the extent to which sera neutralize the antiviral activity of type I IFNs, resulting in efficient infection despite presence of IFNs. We tested all ELISA-positive sera as well as 102 ELISA-negative sera as a reference. 13 (3.2 %) and 8 (2.0 %) of the 403 sera of the CSC specifically neutralized exogenous IFN-□ and IFN-□, respectively, as judged by PCR-based quantification of SARS-CoV-2 genomic RNA in the supernatant and plaque assays that quantify infectivity of virus progeny (Fig. 2a-b, Fig. S1). In cohort D (TPEC), 5 (18.5%) and 3 (11.1%) of 27 sera neutralized IFN-□ and IFN-□ activity, respectively (Fig. 2a-b, Fig. S1).

**Figure 2.**
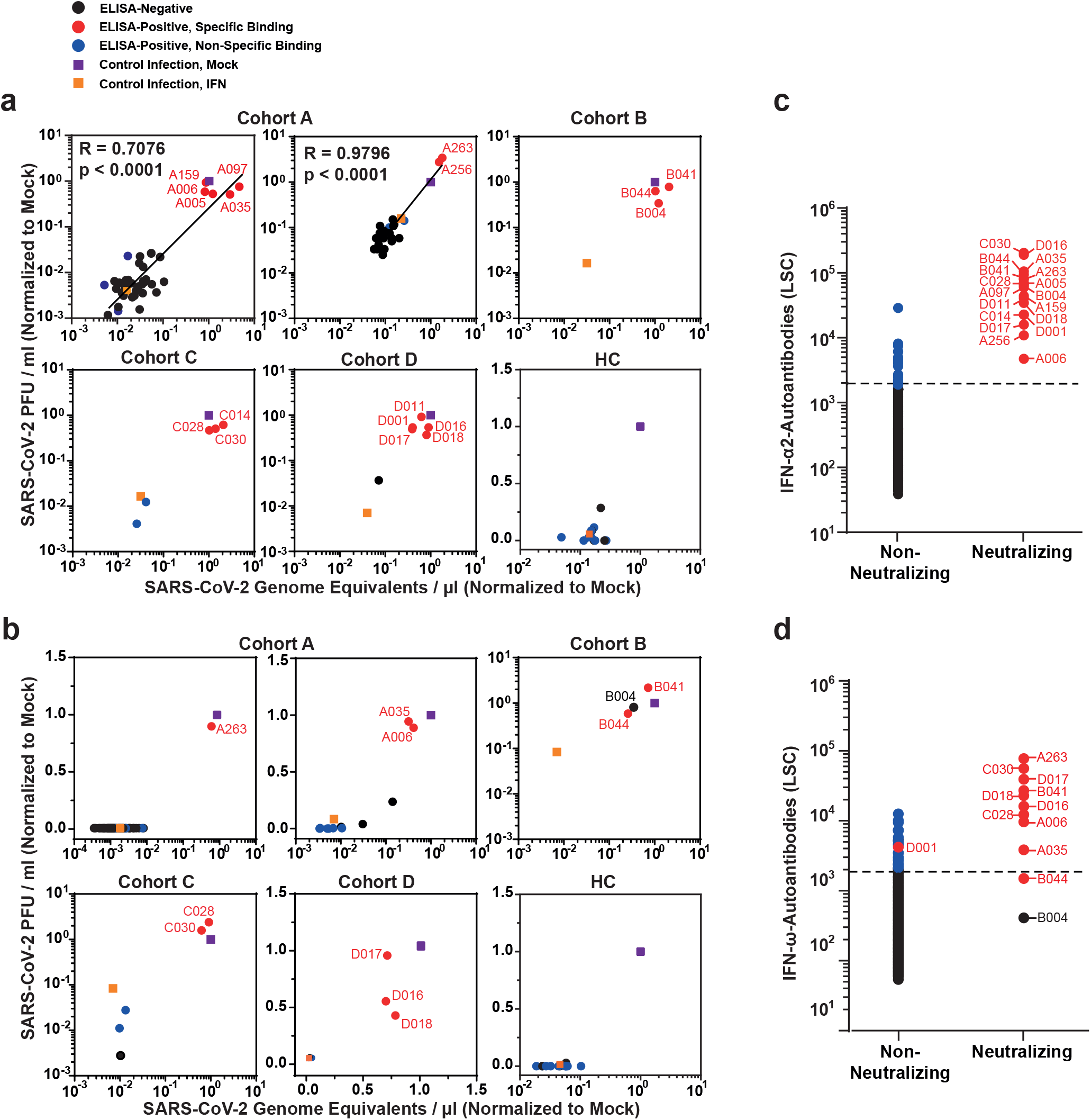
IFN-AABs neutralize exogenous IFN in a virus infection-based assay. (**a**-**b**) Selected sera were analyzed for IFN neutralization activity in a SARS-CoV-2 infection- based assay. The ability of individual sera to neutralize exogenous IFN-α2 (a) and IFN-ω (b) is shown by the rescue of susceptibility to infection as judged by quantification of viral RNA (x-axis) and infectivity (y-axis) in the supernatant. The infection condition in the absence of serum and IFN is set to 1. (**c-d**) The LSC value for individual sera, grouped into non-neutralizing and neutralizing sera. Dots indicate sera containing AABs scoring specific (red) or unspecific (blue) for IFN-α2 and IFN-ω binding in the competition assay (see b), respectively. Black dots indicate samples that scored below the threshold of the ELISA. Black dotted lines indicate the 97.5th percentile of the ECLIA assay LSC in sera from the healthy health care workers (HC) cohort (see Fig. 1). Neutralization ability of IFN-α and IFN-ω can be predicted at 100% for sera displaying LSCs above the respective red dotted lines (IFN-a: 35,639; IFN-ω: 12,603).

Strikingly, among competition assay-positive samples, 18 (100%) of 18, and 9 (90%) of 10 sera displayed IFN- and IFN- -neutralizing activity, respectively, indicating that a positive result in the competition assay associates with neutralization activity with a high likelihood (Fig. 1c-d). Conversely, a negative result in the competition assay was predictive of absence of neutralization ability. Specifically, among ELISA-positive, but competition assay-negative sera, 0 (0%) of 23 and 0 (0%) of 29 sera were able to neutralize IFN-□ and IFN-□, respectively. Among IFN-α AAB-ELISA-negative sera, we observed no IFN-neutralizing activity. Interestingly, among IFN-ω AAB-ELISA-negative sera, we identified two samples that neutralized IFN-ω despite scoring negative in the IFN-ω-AAB ELISA (i.e., having LSC counts below the 97.5th percentile cut-off) (Fig. 2b, d). The pronounced ability of these exact two sera to neutralize IFN-α (Fig. 2a, c) was the reason why we included them in the IFN-ω test, and suggests a potential cross-reactivity of IFN-α-AAB with IFN-ω.

Merging results from all three assays (Fig. 2c-d) revealed that an LSC value in the screening ELISA of >35.639 (IFN-□) and >12.603 (IFN-□) predicted specific binding in the competition assay and neutralization ability in the functional assay. Finally, the prevalence of ten individual antiphospholipid-ABs did not differ between patients with and without neutralizing IFN-AABs from cohort A (Fig. S2), suggesting that the presence of AABs is not generally increased in IFN-AAB-positive patients.

### Clinical phenotype of COVID-19 patients displaying type-I IFN-AABs

We next aimed to characterize the clinical phenotype of IFN-neutralizing AAB-positive COVID-19 patients in the CSC at hospital admission, and to identify discriminatory markers that may serve as pre-selection criteria for their early identification and stratification.

Interestingly, there were no statistically significant differences regarding clinical baseline characteristics, including demographic criteria and pre-existing comorbidities between patients with and without IFN-neutralizing AABs in the CSC using univariate analyses (Table 1). Yet, of all patients with available symptom records (cohorts A and C), the proportion of patients who reported fever and required supplemental oxygen therapy within 72 hours from admission was higher in patients with neutralizing IFN-AABs than in those without (fever: 100%, 12/12 versus 62.6% (234/374), p=0.0079 and oxygen: 100% (12/12) versus 76.0% (250/329), p=0.0528).

Furthermore, patients with IFN-AABs for which respective data were available displayed higher median values of C-reactive protein (CRP), procalcitonin, lactate dehydrogenase (LDH), ferritin, total leukocyte and neutrophil count, as well as neutrophil-to-lymphocyte ratio within the first three days of hospital admission compared to patients without IFN-AABs (Fig. 3, Fig. S3). In addition, patients with neutralizing IFN-AABs showed low levels of CD169/Siglec-1 expression on monocytes, a well-known type I IFN-response marker (Fig. S4). Interestingly, there was a tendency towards a negative correlation between CRP levels and CD169/Siglec-1 within 72 hours from hospital admission.

**Figure 3:**
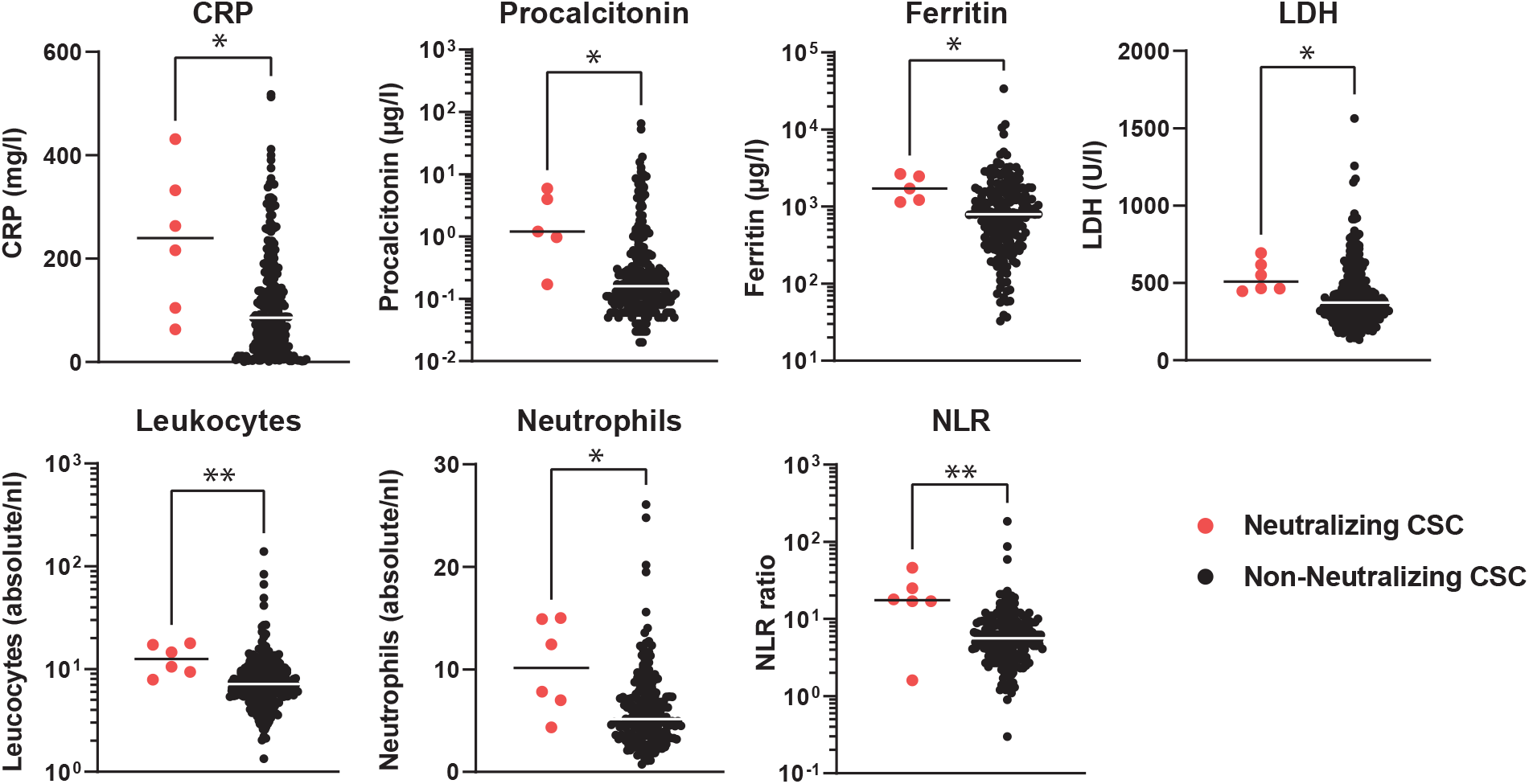
Clinical phenotype of COVID-19 patients displaying type-I IFN-AABs. Values of C-reactive protein (CRP), procalcitonin, ferritin, lactate dehydrogenase (LDH), absolute leukocyte and neutrophil count and neutrophil-to-lymphocyte ratio (NLR) of patients with (N=5-6) and without neutralizing IFN-AABs (N=200-265) from the cross-sectional cohort (CSC, all WHO scores). For each patient, the first available parameter within 72 hours of hospital admission is shown. Statistical testing was performed with Mann-Whitney *U* test.

### In IFN-AAB-positive patients, high quantities of neutralizing IFN-α2-AABs were present both soon post-symptom onset and at the peak of disease

Next, we evaluated the temporal dynamics of IFN-AAB levels in sera from COVID-19 patients soon after symptom onset as compared to the peak of the disease. Available samples obtained in cohort A, and in cohort D (TPEC) prior to TPE were analyzed (Fig. 4, Fig. S5). In all patient sera with detectable neutralizing IFN-AABs at the peak of the disease, early sera corresponding to ten (min. 4 - max. 20) days post-symptom onset (PSO) contained abundant and neutralizing (Fig. 4, Fig. S5) IFN-AABs, suggesting that IFN-AABs either existed prior to the infection or were generated very early PSO.

**Figure 4.**
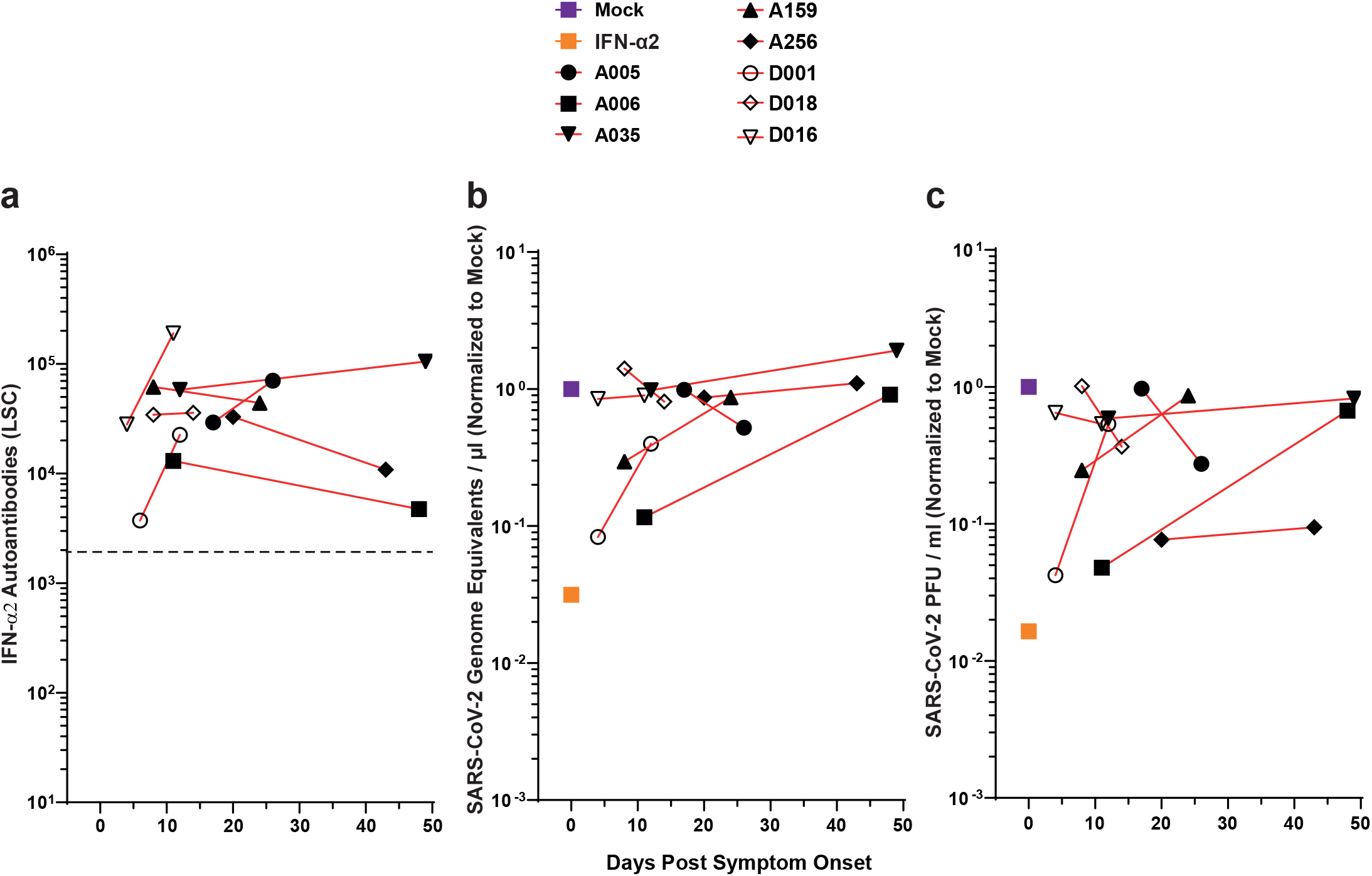
In IFN-AAB-positive patients, high quantities of neutralizing IFN-α2-AABs were present both soon PSO and at the peak of disease. (**a**) Time course of antibody quantities in patient sera that scored IFN-AAB positive at the peak of disease (N=8). The dotted line indicates the 97.5th percentile of the ECLIA assay LSC in sera from the HC cohort (see Fig. 1). (**b-c**) The ability of the sera to neutralize exogenous IFN-α2 is shown by the rescue of susceptibility to infection as judged by quantification of viral RNA (b) and infectivity (c) in the supernatant. The infection condition in the absence of serum and IFN is set to 1.

### Cytokine and humoral responses to SARS-CoV-2 infection in IFN-AAB-positive patients

We next aimed to identify potential quantitative and/or qualitative differences in cytokine responses, viral load and seroconversion kinetics in IFN-AAB-positive as opposed to IFN- AAB-negative patients. We analyzed serum cytokine levels in a subset of critical patients (WHO max. 6-8) from cohort A. Patients with neutralizing IFN-AABs demonstrated significantly higher levels of IFN-γ, and IFN-γ-induced protein 10 (IP-10) one to two weeks PSO while monocyte chemoattractant protein-1 (MCP-1) and TNF-α concentrations were similar compared to sera from patients without neutralizing IFN-AABs (Fig. S6). However, levels equalized among the two groups at three to four weeks PSO. As expected, patients with IFN-AABs had undetectable serum IFN- levels. Of note, by comparing upper-respiratory tract swabs and sera from patients with and without IFN-AABs from all infected cohorts, we failed to identify detectable differences in viral load level or decay over time (Fig. S7a) and we found no evidence for a difference in duration until seroconversion PSO (Fig. S7b). In conclusion, some cytokine responses were aberrantly elevated in patients with IFN- AABs within the first two weeks PSO. However, they normalized at weeks three and four, and viral RNA production and time to seroconversion remained indistinguishable from patients without IFN-AABs.

### Clinical Outcome of COVID-19 Patients with Neutralizing IFN-AABs

Neutralizing IFN-AAB-positive patients developed significantly higher max. WHO scores than patients without neutralizing IFN-AABs (median max. WHO score 8 (IQR 8-8) vs 6 (IQR 4-7), respectively; p<0.0001, Fig. 5a). All patients with neutralizing IFN-AABs in the CSC required IMV (13/13, 100%), compared to 44.5% (166/372) in patients without IFN-AABs (p<0.0001, Fig. 5b). Similarly, the proportion of neutralizing IFN-AAB-positive patients requiring haemodialysis and/or ECMO was markedly higher than in those without IFN-AABs (haemodialysis: 69.2%, 9/13 versus 26.6%, 98/369, p = 0.0008, ECMO: 46.2%, 6/13 versus 15.6%, 58/372, p = 0.0036, Table 1). Twelve out of thirteen neutralizing IFN-AAB-positive patients (92.3%) died in hospital compared to 19.1% (71/372) of patients without IFN-AABs (p<0.0001) in the CSC (Table 1). Median survival of patients with neutralizing IFN-AABs was 28 days (IQR 22-65 days). The probability of surviving to 150 days PSO is 81.3% (300/369) for the patients from the non-neutralizing group, as opposed to 7.7% (1/13) for the patients of the neutralizing IFN-AAB-positive group (Fig. 5c). Conclusively, IFN-AAB positivity was associated with severe disease trajectories of COVID-19 and a worse clinical outcome in our cohorts.

**Figure 5:**
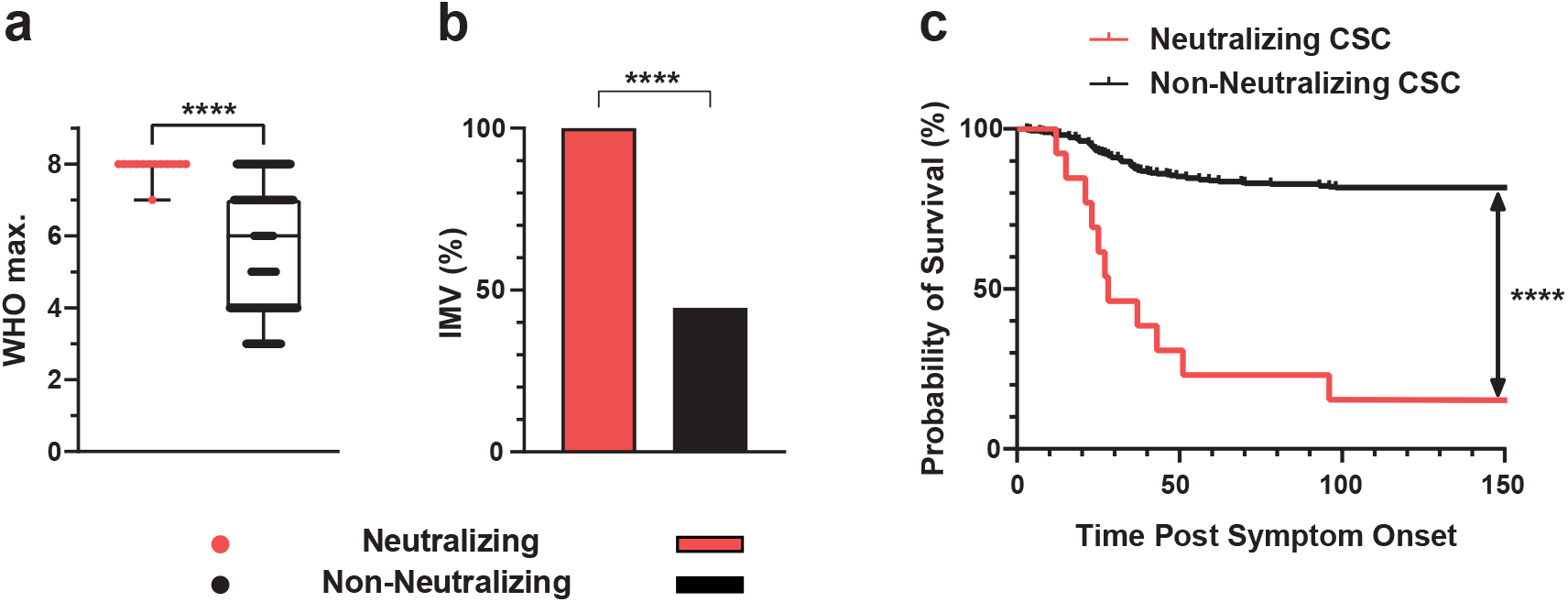
Clinical outcome of COVID-19 patients with neutralizing IFN-AABs. (**a**) Median max. WHO score in hospital. Statistical testing was performed using the Mann­Whitney *U* test. (b) Proportion of patients requiring invasive mechanical ventilation (IMV) after hospital admission. Statistical testing was performed using the Chi-Square test. (c) Probability of survival of patients with and without neutralizing IFN-AABs from the cross­sectional cohort (CSC) from symptom onset until discharge (up to 150 days), death or transferral (p<0.0001). Statistical testing was performed using a log-rank test. Neutralizing (N=13), non-neutralizing (N=372).

### Inter-individual effect of therapeutic plasma exchange on IFN-AABs and SARS-CoV-2 antibodies

Cohort D (TPEC) allowed us to compare trajectories of IFN-AAB-positive patients undergoing TPE to those not undergoing TPE. Criteria for initiation of TPE were: presence of ARDS requiring IMV and/or vasopressor-dependent circulatory shock, clinical and laboratory features of a COVID-19 associated immunopathology with elevated D-dimers and ferritin levels, persistent and refractory fever ≥ 38.5°C without conclusive pathogenic evidence and despite anti-infectious treatment. TPE was first initiated without prior screening for IFN-AABs within a median of six days (IQR 1-10) after hospital admission and the median number of TPE sessions per patient was 3 (IQR 2-5). TPE was performed using a continuous-flow centrifugation blood cell separator. Plasma with enclosed cytokines and immunoglobulins are separated from blood cells by gravity due to different densities of the respective blood components (Nusshag et al. 2021).

Focusing on patients of the WHO group 6-8, survival of IFN-AAB-negative patients in the CSC cohorts and TPEC was similar (p = 0.34). The proportion of neutralizing IFN-AAB-positive patients from the TPEC that survived in hospital was higher than of those patients from the CSC who did not undergo TPE (60%, 3/5 patients from the TPEC survived versus 7.7%, 1/13 patients from the CSC, p=0.0412) (Fig. 6a).

**Figure 6.**
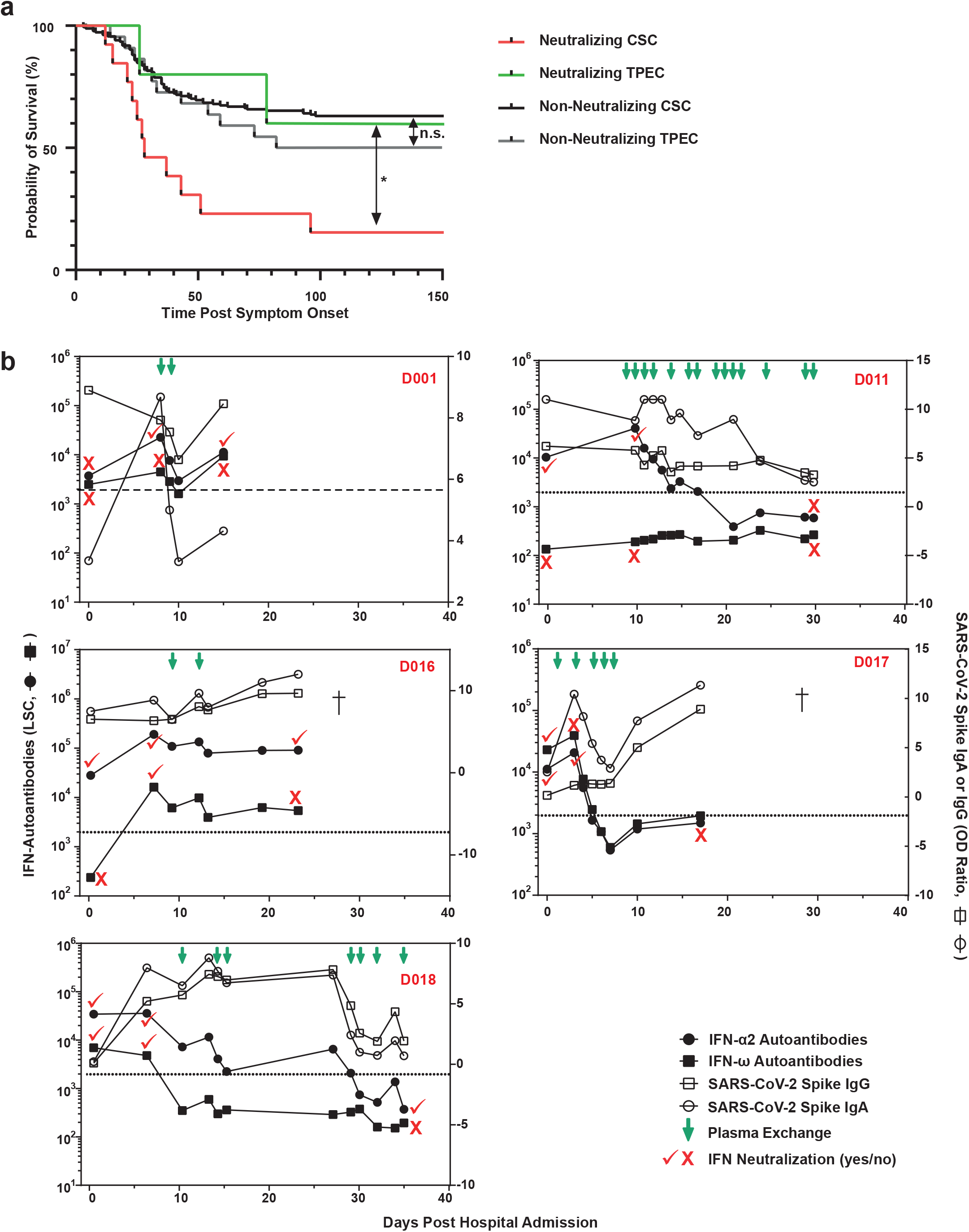
Inter-individual effect of therapeutic plasma exchange on IFN-AABs and SARS-CoV-2 antibodies. (a) Probability of survival of neutralizing IFN-AAB-positive and -negative patients with critical COVID-19 (max. WHO score 6-8) with and without plasma exchange (CSC and TPEC) from symptom onset until discharge, death or transferral (p=0.04, neutralizing CSC versus neutralizing TPEC). Statistical testing was performed using a log-rank test. Neutralizing CSC (N=13), non-neutralizing CSC (N=184), neutralizing TEPC (N=5) and non-neutralizing TPEC (N=22). (b) Antibody profile in serum from individual COVID-19 patients of the TPEC subjected to plasma exchange. The quantity of IFN-α2- and IFN-ω-AABs, SARS-CoV-2-IgG and -IgA, and the IFN-α2 and IFN-ω neutralization status are given for various time points. The patient identifier is given in red. Viral load profiles were only available for patients D011 and D018 and are shown in Supplementary Figure 7.

Longitudinal analysis of sera revealed that three (D011, D017, D018) out of five patients responded to TPE with decreasing IFN-AAB levels below the cut-off and to a level that coincided with absence of neutralizing activity. In addition, a sustained reduction of IFN- AAB quantities was achieved only by repetitive TPE (Fig. 5b, Fig. S8). In contrast to IFN- AABs, quantities of SARS-CoV-2 Spike IgG and IgA were less, if at all, affected by TPE (note the logarithmic scale for IFN-AABs versus the linear scale for SARS-CoV-2-IgG/IgA). Overall, our findings in a limited number of patients suggest that TPE could positively affect the survival of critically ill IFN-AAB-positive patients. This needs to be corroborated in future, adequately powered clinical investigations. Potentially, a sustained and significant reduction of peripheral IFN-AAB levels must be achieved to prevent death.

### Diagnostic algorithm for rapid identification of neutralizing IFN-AAB-positive patients

Given that IFN-AABs are risk factors for a worse clinical outcome in hospitalized patients with COVID-19, future rapid identification of IFN-AAB-positive patients after hospital admission seems key for the potential implementation and success of specific interventions such as TPE. In the CSC cohort, the patient number needed to screen (NNS) without preselection in order to identify one patient with neutralizing IFN-AAB was 31.0 (403/13). We hypothesized that applying clinical pre-selection criteria which strongly co-present with the neutralizing IFN-AAB positivity diminishes the NNS. Using univariate analyses, we established that temperature (>38.5 °C or self-reported) before or upon hospital admission and the need for supplemental oxygen within the first 72 hours after admission correlated best with presence of IFN-AAB positivity in the screening assay in all hospitalized COVID-19 patients (fever: p=0.0079; supplemental oxygen p=0.0528). Using fever and need for supplemental oxygen, the NNS was reduced to 15.6 (172/11). Selection of patients exceeding the cut-off for ELISA positivity (in our cohort 97.5th percentile of the HC) for further testing by competition assay would adjust the NNS in the competition assay to 1.4 (15/11). Therefore, in order to increase sensitivity, we propose to consider the need for supplemental oxygen within 72 hours after admission and fever as pre-selection criteria for patients that undergo ELISA screening (Fig. 7).

**Figure 7.**
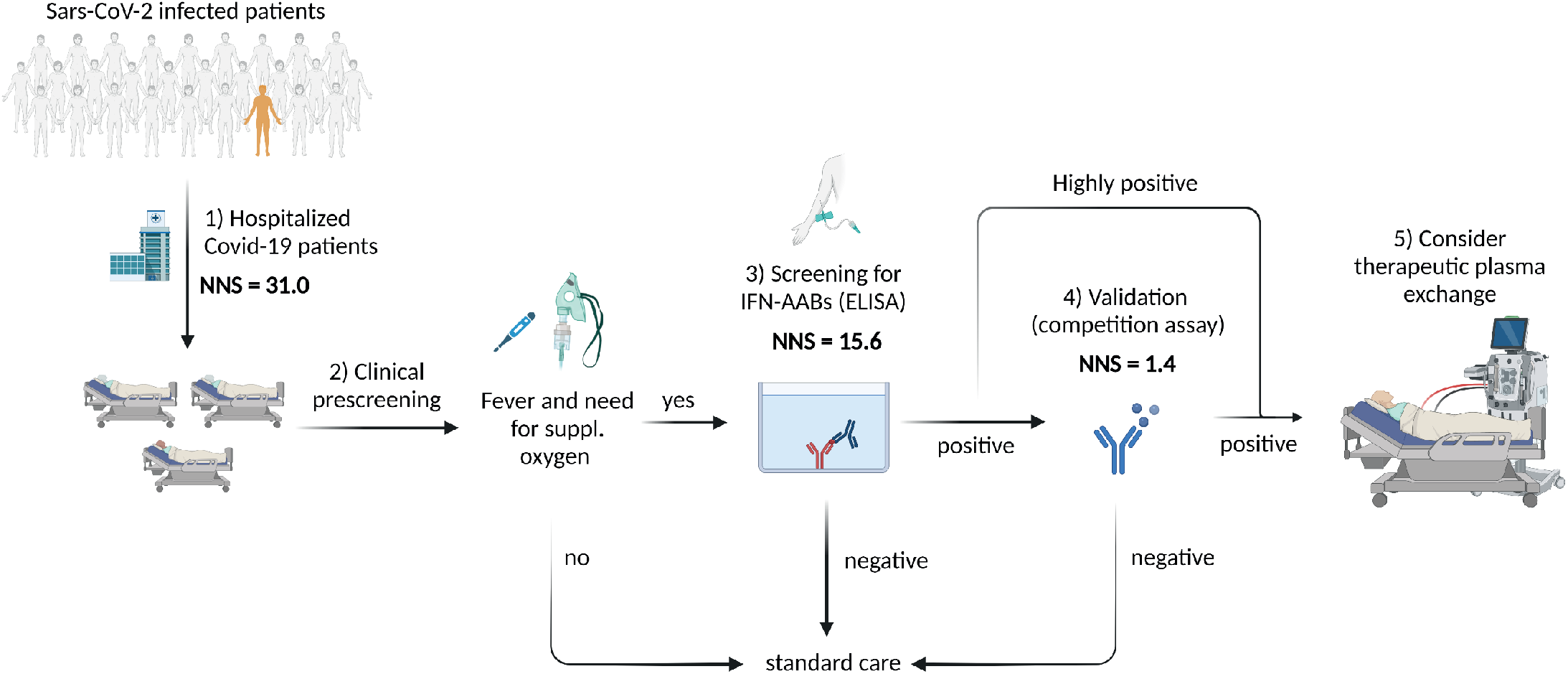
Diagnostic algorithm for rapid identification of neutralizing IFN-AAB-positive patients. The number needed to screen (NNS) is based on results from the cross-sectional cohort (CSC). ELISA for IFN-AAB detection was considered to be positive if it exceeded the 97.5th percentile of the healthy control cohort. (1) NNS of all hospitalized Covid-19 patients without preselection was 31.0 (403 patients in total, 13 patients with neutralizing IFN-AABs). (2) Prescreening of patients using the clinical criteria of fever at admission and need for supplemental oxygen within the first 72 hours after hospitalization diminished the NNS in the IFN-AAB ELISA (3) by half, to 15.6 (172/11). For patients identified as positive in the screening ELISA, the NNS in the competition assay to confirm the presence of IFN-specific AABs is reduced to 1.4 (15/11) (4). For patients with high-titer IFN-AABs (light signal count > 35.639), the competition assay can be omitted. For patients highly positive in the IFN-AAB ELISA and those with specific results in the competition assay, we propose to consider therapeutic plasma exchange within clinical studies (5).

Sera from all patients exceeding the LSC value of 35,639 (13/13) in the screening ELISA assay demonstrated neutralizing activity against IFN-α (Fig. 2c). We therefore propose to conduct the IFN neutralization assay only in case of an LSC value lower than 35,639, whereas patients with sera exceeding this value can be considered positive for neutralizing IFN-AABs without further testing (Fig. 7).

## DISCUSSION

IFN-AABs strongly associate with adverse clinical outcome of SARS-CoV-2 infection (Bastard et al. 2020; Koning et al. 2021; Troya et al. 2021; Goncalves et al. 2021; Bastard et al. 2021; Lemarquis et al. 2021; de Prost et al. 2021). Unfortunately, the inability to identify IFN-AAB-positive COVID-19 patients early enough before deterioration delays their access to specific, yet untested therapies that may require implementation directly upon hospital admission. In several studies, detection and quantification of IFN-AABs in sera from COVID- 19 patients relies on ELISA and multiplex particle-based assay. While these assays are amenable to high-throughput and are highly sensitive, they result in a small proportion of false-positive results (Güven et al. 2014; Sahud et al. 2012), highlighting the ongoing need to reanalyze positive-tested patient material in functional assays demonstrating the neutralization activity. However, such assays are sophisticated and time-consuming. They include luciferase-based interferon-stimulated response element (ISRE) promoter reporter assays (Troya et al. 2021; Goncalves et al. 2021), flow cytometry-based analyses of STAT phosphorylation (Bastard et al. 2020; Goncalves et al. 2021; Meisel et al. 2021), and virus infection-based assays (de Prost et al. 2021; Meisel et al. 2021). The latter allows probing the activity of the IFN-AABs in the context of infection-inhibitory concentrations of IFN-α and IFN-ω. Here, we applied and cross-validated previously established assays comprising an ELISA for sensitive identification, a specificity-validating competition assay, and a functional neutralization assay (Meisel et al. 2021) using a large collection of serum samples obtained from three cross-sectional cohorts. Equally important, we identified clinical parameters that co-present with IFN-AAB positivity at hospital admission. Combination of clinical parameters and targeted diagnostic testing results in an algorithm for early, sensitive, specific identification of IFN-AAB-positive patients which can be easily introduced into clinical routine diagnostics.

Surprisingly, sera from two patients were found to neutralize exogenous IFN-ω despite negative ELISA results. Presence of neutralization activity in the absence of detectable IFN-AABs has been reported (Bastard et al. 2021). Explanations for this phenomenon could include technical aspects of the detection method, including the possibility that IFN-AABs may be concealed by the binding of the cytokine to the plate or biotinylation of the cytokine (Meyer et al. 2016).

Here, we calibrated our ELISA cut-off based on the 97.5th percentile in a cohort of uninfected individuals. Although this strategy may be inexact, the absence of prevalence IFN-AABs in a cohort of younger and predominantly female healthcare workers supports an age-dependent increase of IFN-AAB prevalence in uninfected individuals (Bastard et al. 2021). Furthermore, the prevalence of 3.2% (13/403) of patients with neutralizing AABs against IFN-α and/or IFN-ω in our cross-sectional patient cohort (median max. WHO score 6) and 18.5% (5/27) in critically affected patients (median max. WHO-Score 7) is in line with reported prevalences of 6-17% in severely (Bastard et al. 2020; Koning et al. 2021; Troya et al. 2021; van der Wijst et al. 2021) and 11-19% in critically ill (Bastard et al. 2020; van der Wijst et al. 2021; Goncalves et al. 2021) individuals with COVID-19. Moreover, IFN-AAB positivity is associated with a worse clinical outcome and a decreased survival probability of hospitalized patients in our cohorts, confirming previous reports (Bastard et al. 2020; Koning et al. 2021; Troya et al. 2021; Goncalves et al. 2021; Bastard et al. 2021; Lemarquis et al. 2021; de Prost et al. 2021). Interestingly, a single study to date (Abers et al. 2021) suggested that survival was not adversely affected by the presence of type-I IFN-AABs, while confirming the widely accepted association with an increased risk of admission to the intensive care unit.

We failed to identify a clear association of IFN-AABs with previously described demographic parameters in our cross-sectional cohort, including male sex or advanced age (Bastard et al. 2020, 2021), probably due to the relatively limited sample size of our cohorts. However, the presence of neutralizing IFN-AABs was associated with fever and need for supplementary oxygen within 72 hours post hospital admission, as well as with elevated soluble and cellular markers of acute-phase reaction including elevated levels of CRP, procalcitonin, LDH and ferritin and elevated total neutrophil and leukocyte counts within the first three days of admission in our CSC. Higher CRP values constitute a biomarker for a severe disease course, are included in a widely-used clinical risk score for mortality of COVID-19 (Knight et al. 2020), and associate with neutralizing IFN-AABs along with lower lymphocyte counts in severely affected patients (Troya et al. 2021). As hospital admission and thus clinical deterioration occurred at a median of five days PSO in the CSC, we propose fever and need for supplemental oxygen therapy up to 72 hours post hospital admission as suitable and simple clinical criteria to identify patients at risk for a severe disease course.

Our ability to detect IFN-AABs as early as four days PSO in sera from most patients that present with IFN-AABs at the peak of their disease suggest that they were present prior to the infection, or alternatively, but less likely, were induced very early post infection. Our data are in agreement with recently demonstrated presence of IFN-AABs at the day of hospital admission (de Prost et al. 2021) and in 4% of uninfected individuals >70 years old (Bastard et al. 2021), underlining the idea that they can serve as biomarkers for predisposition for a severe course of SARS-CoV-2 infection. Future studies are required to elucidate the biological mechanisms that lead to elicitation of IFN-AABs in an age-dependent manner.

Currently discussed therapeutic options tailored to IFN-AAB-positive COVID-19 patients comprise mainly TPE and IFN-β administration. TPE in the context of COVID-19 has been analyzed in individual case reports and case-control studies, including IFN-AAB-positive and negative patients (Lemarquis et al. 2021; Nusshag et al. 2021; de Prost et al. 2021; Fernandez et al. 2020) and might efficiently remove soluble circulating Fc gamma receptor-activating immune complexes (Ankerhold et al. 2021). TPE effectively decreased circulating IFN-AAB, but not SARS-CoV-2 antibody concentrations in four IFN-AAB-positive, severely ill patients (de Prost et al. 2021) and in a child with APS-1 suffering from severe COVID-19 (Lemarquis et al. 2021). In our study, TPE reduced circulating IFN-AABs with patient-specific efficiency and appeared to increase the chances of in-hospital survival, underlining the rationale to initiate large-scale, adequately powered clinical trials in order to corroborate the potential benefit of TPE in a general cohort of adult, critically ill COVID-19 patients. Interestingly, SARS-CoV-2-IgG and IgA quantities were less affected by TPE for unknown reasons, which may include their rapid replenishment by highly abundant plasmablasts or an extravascular-to-intravascular rebound since immunoglobulins have a substantial extravascular distribution.

Given the reported absence of detectable IFN-β-AABs, IFN-β administration may substitute neutralized IFN-α and -ω. While IFN-β therapy failed to result in a detectable clinical benefit in the SOLIDARITY trial (WHO Solidarity Trial Consortium et al. 2021), specifically IFN-AAB-positive patients may benefit from IFN-β therapy, a patient group that might have been under-represented in this study. Furthermore, the benefit of IFN-β administered by different routes should be systematically explored in this patient group.

## CONCLUSIONS

Generation of comprehensive data on the clinical benefit of TPE and IFN-β therapy are hampered by the difficulty of identifying IFN-AAB-positive patients for enrollment in large clinical studies. Our diagnostic algorithm for their identification may support clinicians to promptly identify patients carrying this additional risk factor for progression to severe disease upon hospital admission. It is based on clinical preselection of patients presenting with fever and need for supplemental oxygen therapy within 72 hours after admission to be further screened by an ELISA-based assay for circulating IFN-AABs and an easily implementable

ELISA-based validation assay which may be skipped in case of high positivity in the screening assay (Fig. 7). Screening criteria should be validated in the future in a large prospective observational cohort. Ideally, methodological complex gold-standard assays can be substituted by high-throughput diagnostic tests and stratification by clinical parameters, facilitating clinical implementation. Our algorithm may allow identifying at-risk patients upon presentation in the emergency room to be directly allocated to TPE and/or IFN-β therapy within larger clinical trials, which are urgently needed to determine clinical effectiveness of these targeted therapies.

## Data Availability

All data produced in the present study are available upon reasonable request to the authors

## Acknowledgements

We thank Mahtab Maleki for support with IFN-AAB ELISAs and competition assays. We thank Patricia Tscheak, Marie Luisa Schmidt, and Tatjana Schwarz for support with sample logistics and SARS-CoV-2 antibody testing. V.M.C is a participant of the Charité Clinician Scientist program funded by Charité - Universitätsmedizin Berlin and the Berlin Institute of Health. We thank Karin Reiter for the anti-phospholipid antibody measurements. We thank the patients for their ongoing trust and collaboration. We thank the involved biobanks (ZeBanC Berlin, FREEZE-Biobank Freiburg) for their support. DD and AL are members of SFB1425, funded by the Deutsche Forschungsgemeinschaft (DFG, German Research Foundation) - Project #422681845. NS is partly funded by the Deutsche Forschungsgemeinschaft (DFG, Research Grant No. SCHA1838/4-2).

## Pa-COVID study Group (Consortium representative: Florian Kurth)

### Set up study platform

Stefan Hippenstiel^4^, Martin Witzenrath^4^, Elisa T. Helbig^4^, Lena J. Lippert^4^, Pinkus Tober-Lau^4^, David Hillus^4^, Sarah Steinbrecher^4^, Sascha S. Haenel^4^, Alexandra Horn^4^, Willi M^4^. Koch, Nadine Olk^4^, Rosa C. Schuhmacher^4^, Katrin K. Stoyanova^4^, Lisa Ruby^4^, Claudia Zensen^4^, Mirja Mittermaier^4^, Fridolin Steinbeis^4^, Tilman Lingscheid^4^, Bettina Temmesfeld-Wollbrück^4^, Thomas Zoller^4^, Holger Müller-Redetzky^4^, Alexander Uhrig^4^, Daniel Grund^4^, Christoph Ruwwe-Glösenkamp^4^, Miriam S. Stegemann^4^, Katrin M. Heim^4^, Bastian Opitz^4^, Kai-Uwe Eckardt^26^, Martin Möckel^27^, Felix Balzer^28^, Claudia Spies^28^, Steffen Weber-Carstens^28^, Frank Tacke^29^, Chantip Dang-Heine^2^, Michael Hummel^30^, Georg Schwanitz^31^, Uwe D. Behrens^31^, Maria Rönnefarth^2^, Sein Schmidt^2^, Alexander Krannich^2^, Saskia Zvorc^2^, Jenny Kollek^2^ and Christof von Kalle^2^.

### Inclusion of patients and clinical data curation

Linda Jürgens^4^, Malte Kleinschmidt^4^, Sophy Denker^32^, Moritz Pfeiffer^4^, Belén Millet Pascual-Leone^4^, Luisa Mrziglod^4^, Felix Machleidt^4^, Sebastian Albus^4^, Felix Bremer^4^, Tim Andermann^4^, Carmen Garcia^4^, Philipp Knape^4^, Philipp M. Krause^4^, Liron Lechtenberg^4^, Yaosi Li^4^, Panagiotis Pergantis^4^, Till Jacobi^4^, Teresa Ritter^26^, Berna Yedikat^4^, Lennart Pfannkuch^4^, Ute Kellermann^4^, Susanne Fieberg^4^, Laure Bosquillon de Jarcy^1^, Anne Wetzel^4^, Markus C. Brack^4^, Moritz Müller-Plathe^4^, Jan M. Kruse^26^, Daniel Zickler^26^, Andreas Edel^28^, Britta Stier^26^, Roland Körner^26^, Nils B. Müller^26^, Philipp Enghard^26^, Lucie Kretzler^2^, Lil A. Meyer-Arndt^33^, Linna Li^2^, and Isabelle Wirsching^2^

### Biobanking and -sampling

Denise Treue^30^, Dana Briesemeister^30^, Jenny Schlesinger^30^, Birgit Sawitzki^25^, Lara Bardtke^4^, Kai Pohl^4^, Philipp Georg^4^, Daniel Wendisch^4^, Anna L. Hiller^4^, Sophie Brumhard^4^, Marie Luisa Schmidt^1^, Leonie Meiners^1^, and Patricia Tscheak^1^

## Study Group-Specific Affiliations

^26^Charité - Universitätsmedizin Berlin, corporate member of Freie Universität Berlin and Humboldt-Universität zu Berlin, Institute of Nephrology and Internal Intensive Care Medicine, Berlin, Germany

^27^Charité - Universitätsmedizin Berlin, corporate member of Freie Universität Berlin and Humboldt-Universität zu Berlin, Division of Emergency Medicine and Department of Cardiology, Berlin, Germany

^28^Charité - Universitätsmedizin Berlin, corporate member of Freie Universität Berlin and Humboldt-Universität zu Berlin, Department of Anesthesiology and Intensive Care Medicine, Berlin, Germany

^29^Charité - Universitätsmedizin Berlin, corporate member of Freie Universität Berlin and Humboldt-Universität zu Berlin, Department of Hepatology and Gastroenterology, Berlin, Germany

^30^Charité - Universitätsmedizin Berlin, corporate member of Freie Universität Berlin and Humboldt-Universität zu Berlin, Central Biobank Charité (ZeBanC), Institute of Pathology, Berlin, Germany

^31^Charité - Universitätsmedizin Berlin, corporate member of Freie Universität Berlin and Humboldt-Universität zu Berlin, Clinical Study Center, Berlin, Germany

^32^Charité - Universitätsmedizin Berlin, corporate member of Freie Universität Berlin and Humboldt-Universität zu Berlin, Department of Hematology, Oncology and Tumor Immunology, Berlin, Germany

^33^Charité - Universitätsmedizin Berlin, corporate member of Freie Universität Berlin and Humboldt-Universität zu Berlin, Department of Neurology, Berlin, Germany

## Author contributions

*Conceptualization*: Florian Kurth; Horst von Bernuth; Christian Meisel; Christine Goffinet *Methodology*: Bengisu Akbil; Tim Meyer; Olga Staudacher

*Formal analysis and investigation*: Bengisu Akbil; Tim Meyer; Paula Stubbemann; Charlotte Thibeault; Olga Staudacher; Jenny Jansen; Barbara Mühlebach; Jan-Moritz Doehn; Christoph Tabeling; Christian Nusshag, Cédric Hirzel; David Sökler Sanchez; Daniel Duerschmied, Achim Lother; Nils Schallner; Jan Nikolaus Lieberum; Dietrich August; Siegbert Rieg; Valeria Falcone; Uwe Kölsch; Nadine Unterwalder; Thibaud Spinetti; Joerg Schefold; Thomas Dörner; Victor M. Corman; Uta Merle

*Writing - original draft preparation*: Paula Stubbemann; Charlotte Thibeault; Christine Gofffinet

*Writing - review and editing*: Bengisu Akbil; Tim Meyer; Paula Stubbemann; Charlotte Thibeault; Olga Staudacher; Daniela Niemeyer; Barbara Mühlemann; Christian Nusshag; Daniel Duerschmied, Achim Lother; Terry C. Jones; Klaus Warnatz; Thibaud Spinetti; Joerg C. Schefold; Leif Sander; Victor M. Corman; Uta Merle; Florian Kurth; Horst von Bernuth; Christian Meisel; Christine Goffinet Funding acquisition: Joerg Schefold; Klaus Warnatz; Uta Merle; Florian Kurth; Horst von Bernuth; Christian Meisel; Christine Goffinet

*Resources*: Daniela Niemeyer, Alexandra Nieters; Christian Drosten; Leif Sander; Florian Kurth; Horst von Bernuth; Christian Meisel; Christine Goffinet

Supervision: Hartmut Hengel; Ralf-Harto Hübner; Norbert Suttorp; Klaus Warnatz; Victor M. Corman; Florian Kurth; Horst von Bernuth; Christian Meisel; Christine Goffinet

## Funding

This work was supported by the Innovationsfond of Labor Berlin (to H.V.B, C.M., and C.G.); by funding from the Deutsche Forschungsgemeinschaft (DFG) Collaborative Research Centre CRC900 “Microbial Persistence and its Control,” project number 158989968, project C8 (awarded to C.G.); by funding from the Berlin Institute of Health (BIH) (to C.G.). T.C.J. is in part funded through NIAID-NIH CEIRS contract HHSN272201400008C. Parts of the work were funded by the European Union’s Horizon 2020 research and innovation program through project RECOVER (GA101003589) to C.D.; the German Ministry of Research through the projects VARIPath (01KI2021) to V.M.C., and NaFoUniMedCovid19 - COVIM, FKZ: 01KX2021 to C.D., V.M.C., L.E.S., F.K. and H.H. The Pa-COVID-19 Study is funded by BIH and the German Ministry of Research NaFoUniMedCovid19 NUM-NAPKON FKZ: 01KX2021. The COV-IMMUN study is supported by the Berlin University Alliance and the BIH Clinical Study Center (BIH-CSC). KW has received funding by the Deutsche Forschungsgemeinschaft (DFG, Research Grant No. WA 1597/6-1 and WA 1597/7-1).

## Disclosure of Conflicts of interest

V.M.C is named together with Euroimmun GmbH on a patent application filed recently regarding the diagnostic of SARS-CoV-2 by antibody testing. Technische Universität Berlin, Freie Universität Berlin and Charité - Universitätsmedizin have filed a patent application for siRNAs inhibiting SARS-CoV-2 replication with D.N. as co-author. J.C.S, T.S (full departmental disclosure): the department of Intensive Care Medicine has/had research and/or development/consulting contracts with (full disclosure) Orion Corporation, Abbott Nutrition International, B. Braun Medical AG, CSEM SA, Edwards Lifesciences Services GmbH/SA, Kenta Biotech Ltd, Maquet Critical Care AB, Omnicare Clinical Research AG, Phagenesis Ltd, Cytel, and Nestlé. No personal financial gains resulted from respective development/consulting contracts and/or educational grants.

## Research involving human participants

All studies were conducted according to the Declaration of Helsinki and Good Clinical Practice principles (ICH 1996).

The Pa-COVID-19 study was approved by the ethics committee of Charité - Universitätsmedizin Berlin (EA2/066/20) and is registered in the German and WHO international clinical trials registry (DRKS00021688).

The Covimmun-study was approved by the ethics committee of Charité - Universitätsmedizin Berlin (EA1/068/20).

The study “Evaluation der humoralen und zellulären Immunantwort und der thromboembolischen Komplikationen bei Infektion mit dem neuartigen Coronavirus (nCoV19)” was approved by the Kantonale Ethikkommission KEK, Bern, Switzerland, Nr. 2020-00877 and is registered under the name “Characterizing the Immune Response and Neuronal Damage in COVID-19” at http://clinicaltrials.gov (NCT04510012).

The three studies (COVID-19 Register Freiburg; FREEZE-Covid19 cohort and Biomarker COVID-19) were approved by the ethics committee of the University of Freiburg (EK-FR 153/20; FREEZE-Biobank EK-FR 383/19; EK-FR 225/20) and are registered in the German and WHO international clinical trials registry (DRKS00021522 for Biomarker COVID-19).

The Covid-Immun study was approved by the local Ethics Committee of the Medical Faculty of Heidelberg (S148/2020) and is registered in the German clinical trials registry (DRKS00021810).

## Informed consent

Informed consent was obtained from all individual participants included in the studies.

## Availability of data and material

The datasets and materials generated during and/or analyzed during the current study are available from the corresponding authors on reasonable request.

## Code availability

Not applicable. No data were coded in this study.

## Notes

### Author Declarations

All studies were conducted according to the Declaration of Helsinki and Good Clinical Practice principles (ICH 1996). The Pa-COVID-19 study was approved by the ethics committee of Charite Universitaetsmedizin Berlin (EA2/066/20) and is registered in the German and WHO international clinical trials registry (DRKS00021688). The Covimmun-study was approved by the ethics committee of Charite Universitaetsmedizin Berlin (EA1/068/20). The study Evaluation der humoralen und zellulaeren Immunantwort und der thromboembolischen Komplikationen bei Infektion mit dem neuartigen Coronavirus (nCoV19) was approved by the Kantonale Ethikkommission KEK, Bern, Switzerland, Nr. 2020-00877 and is registered under the name Characterizing the Immune Response and Neuronal Damage in COVID-19 at http://clinicaltrials.gov (NCT04510012). The three studies (COVID-19 Register Freiburg; FREEZE-Covid19 cohort and Biomarker COVID-19) were approved by the ethics committee of the University of Freiburg (EK-FR 153/20; FREEZE-Biobank EK-FR 383/19; EK-FR 225/20) and are registered in the German and WHO international clinical trials registry (DRKS00021522 for Biomarker COVID-19). The Covid-Immun study was approved by the local Ethics Committee of the Medical Faculty of Heidelberg (S148/2020) and is registered in the German clinical trials registry (DRKS00021810).

