## Supplemental File for "Early and rapid identification of COVID-19 patients with neutralizing type I-interferon auto-antibodies by an easily implementable algorithm"

### SUPPLEMENTARY INFORMATION

### Supplementary methods

#### Study characteristics cohorts A-D

Inclusion criteria for cohorts A-D were PCR-confirmed SARS-CoV-2 infection and written informed consent by either patient or legal representative. Studies A-D were monocentric, prospective observational studies. Patients were treated and allocated according to structured regional processes. Patients' treatment was unaffected by participation in the study. Management of critically ill patients followed national and international recommendations or was performed as compassionate use, at the discretion of the treating physicians (cohort D).

*Cohort A - Berlin*

In total, 266 adult hospitalized patients were recruited into the Pa-COVID-19 study from March to December, 2020, as described (Kurth et al. 2020). The study was approved by the ethics committee of Charité – Universitätsmedizin Berlin (EA2/066/20) and is registered in the German and WHO international clinical trials registry (DRKS00021688).

*Cohort B - Bern*

In total, 50 adult hospitalized patients were recruited into the study “Evaluation der humoralen und zellulären Immunantwort und der thromboembolischen Komplikationen bei Infektion mit dem neuartigen Coronavirus (nCoV19)” from March until October 2020, as described earlier (Spinetti et al. 2020). The study was approved by the Kantonale Ethikkommission KEK, Bern, Switzerland, Nr. 2020-00877 and is registered at http://clinicaltrials.gov (NCT04510012).

*Cohort C - Freiburg*

In total, 87 adult hospitalized patients were enrolled into one of three studies conducted at University Medical Center Freiburg (COVID-19 Register Freiburg; FREEZE-Covid19 cohort; Biomarker COVID-19) from March 2020 until March 2021. The studies were approved by the ethics committee of the University of Freiburg (EK-FR 153/20; FREEZE-Biobank EK-FR 383/19; EK-FR 225/20) and are registered in the German and WHO international clinical trials registry (DRKS00021522 for Biomarker COVID-19).

*Cohort D - Heidelberg*

All adult hospitalized COVID-19 patients treated at the Department of Internal Medicine IV of the University Hospital Heidelberg were enrolled (recruitment start March 18$^{th}$, recruitment ongoing). Of these, 27 who underwent TPE as compassionate use were retrospectively selected for the TPEC and included in this analysis. TPE was performed using a continuous-flow centrifugation blood cell separator (Comtech, Fresenius Medical Care AG & Co. KGaA, Bad Homburg, Germany) centrifuge against fresh frozen plasma (FFP), exchanging a plasma volume of 50 mL per kilogram of body weight or a maximum of 4000 mL per treatment. According to the center standard, all patients with ARDS were empirically treated with Piperacillin-Tazobactam and received a prophylactic antifungal therapy with Caspofungin.

Anti-infective therapy was adjusted according to microbiological evidence. The study was approved by the local Ethics Committee of the Medical Faculty of Heidelberg (S148/2020, DRKS00021810).

**Data collection**

To assess disease severity, the WHO ordinal scale for clinical improvement was used, ranging from 3 to 8 (3 = hospitalized without need of oxygen therapy, 4 = hospitalized with need of oxygen therapy, 5 = high-flow oxygen therapy, 6 = invasive mechanical ventilation without organ supplementation, 7 = invasive mechanical ventilation with organ supplementation, 8 = death). Day of symptom onset was documented according to self-reporting of patients or close contact persons. Fever was defined as measured body temperature of ≥ 38.5°C in hospital or self-reporting of fever by either patient or close contact person PSO. All routine laboratory parameters were measured in accredited laboratories at the respective study site.

*Health care workers*

Six-hundred-and-sixty-seven health-care workers were enrolled into the Covimmun-study (Schwarz et al. 2021), a prospective observational cohort study between March and September, 2020. Inclusion criteria were employment at Charité – Universitätsmedizin Berlin and written informed consent. The study was approved by the ethics committee of Charité – Universitätsmedizin Berlin (EA1/068/20).

**Sample selection**

After centrifugation at room temperature, the undiluted serum was stored in aliquots at -80°C prior to further use. For the HC, we analyzed serum samples obtained at day of inclusion. For screening and validation assays in cohorts A-D, we analyzed sera obtained at the peak of the disease, i.e., week 3-4 PSO. Where indicated, we included sera collected longitudinally, including earlier time points of the disease.

**Cell culture**

Calu-3 and Vero E6 cells were cultured in DMEM (Dulbecco's Modified Eagle's Medium, Sigma Aldrich) with 4.5 g/L glucose, supplemented with 10% fetal bovine serum, 1% penicillin/streptomycin, and 1% L-glutamine at 37 °C and 5% $CO_2$.

**Virus infection-based neutralization assays**

SARS-CoV-2 B.1 lineage virus (strain: BetaCoV/Munich/ChVir984/2020, , EPI_ISL_406862; (Wölfel et al. 2020)) was propagated in Vero E6 cells and sequence integrity was verified by next-generation sequencing. Calu-3 cells were seeded at a density of 6 x $10^5$ cells/ml and pre-incubated with 1% human serum in the presence or absence of 200-400 IU/ml IFN-α2a (Roferon®-A, Roche) or 20-50 ng/ml IFN-ω (PeproTech). After 24 hours, IFN and serum were removed and cells were infected with SARS-CoV-2 at MOI 0.01. Virus inoculum was removed after one hour, cells were washed with PBS, and 100 µl medium were added per well. 24 hours post infection, cell culture supernatant was collected for viral RNA quantification and infectious titer determination. For viral RNA extraction, 50 µl of cell culture supernatant were mixed with 300 µl MagNA Pure external lysis buffer (Roche, Penzberg, Germany) and heat-inactivated at 70°C for ten minutes. Extraction was performed by automated pipetting using MagNA Pure 96 instrument (Roche). For plaque assays, 30 µl of cell culture supernatant were mixed with 30 µl of 0.5% gelatin in Opti-Pro™ for stabilization of infectious particles during storage at -80°C. All SARS-CoV-2-related infection experiments were performed under biosafety level-3 (BSL-3) conditions with enhanced respiratory personal protection equipment.

*Real-time reverse-transcription PCR*

SARS-CoV-2 RNA concentrations were quantified by real-time RT-PCR assay targeting the SARS-CoV-2 E gene using the following primers (Corman et al. 2020): E_Sarbeco_F: ACAGGTACGTTAATAGTTAATAGCGT; E_Sarbeco_P1: FAM-ACACTAGCCATCCTTACTGCGCTTCG-BBQ; E_Sarbeco_R:

ATATTGCAGCAGTACGCACACA. Real time RT-PCRs were performed using the Superscript III OneStep RT-PCR kit (Invitrogen, Darmstadt, Germany). Reactions were performed in 12.5 µl volume with 6.25 µl of 2x reaction buffer, 0.5 µl of 10 µM forward and reverse primers, 0.25 µl of 10 µM probe, 0.5 µl of SSIII / P. Taq enzyme mix, RNase-free water, and 2.5 µl of RNA template. RNA was reverse transcribed at 55°C for 10 min, followed by initial denaturation at 95°C for 180 s. Cycling and fluorescence signal acquisition was performed for 45 cycles of 95°C for 15 s and 58°C for 30 s. Real-time RT-PCR experiment and data processing was conducted using the LightCycler® 480 Real-Time PCR System (Roche). Absolute quantification was performed using SARS-CoV-2-specific *in vitro*-transcribed RNA standards (Corman et al. 2020).

*Plaque assay*

Infectious SARS-CoV-2 plaque-forming units (PFU) were quantified by plaque titration. Vero E6 cells were seeded at a density of $3.5 \times 10^5$ cells/ml in 24-well plates and infected with serial dilutions of SARS-CoV-2-containing cell culture supernatants for one hour at 37°C. After removing the inoculum, cells were washed with PBS and overlaid with 2.4% Avicel (FMC BioPolymers) 1:2 diluted in 2× DMEM supplemented with 2% penicillin/streptomycin, 2% L-glutamine, 2% non-essential amino acids, 2% sodium pyruvate and 20% fetal bovine serum. Three days after infection, the overlay was discarded, cells were fixed in 6% formaldehyde and stained with a 0.2% crystal violet, 2% ethanol and 10% formaldehyde-containing solution. PFU/ml were determined by counting the plaques in the respective dilutions.

**Cytokine and chemokine measurements**

For cytokine and chemokine measurements, a subset of critically ill patients from cohort A (WHO max. ≥ 6) with available biosamples once a week within the first two weeks PSO were selected. IFN-ɑ and IL-28 were measured using Quanterix' single molecule array technology (Simoa) and IL-10, IL-12p70, IL-17A, IL-1α, IL-1β, IL-4, IL-22, IP-10, MCP-1, TNF-α and IFNγ were determined by an ECLIA multiplex assay using the Meso Scale Discovery Platform (MSD) according to the manufacturer's instructions.

**Antiphospholipid antibody testing**

A total of 40 sera from center A (all available sera at the peak of the disease (same time point as measurements of IFN-AABs) of patients with neutralizing IFN-AABs (n=6) and 34 samples from hospitalized SARS-CoV-2 positive controls without neutralizing IFN-AABs) were screened for presence of ten different antiphospholipid AABs (Cardiolipin, Phosphatidic acid, Phosphatidylcholine, Phosphatidylethanolamine, Phosphatidylglycerol, Phosphatidylinositol, Phosphatidylserine, Annexin V, beta-2-GPI, and Prothrombin) using the anti-phospholipid 10 dot LINE Immunoassay (LIA; Generic Assay GmbH Dahlewitz, Berlin) according to the manufacturer's instruction for the determination of IgG and IgM antibodies.

**Serology against SARS-CoV-2**

Samples obtained within clinical routine and weekly as part of the study protocol (Center A) were examined for presence of SARS-CoV-2 specific antibodies (IgG and IgA) to the spike protein using an ELISA kit (Euroimmun, Lübeck, Germany) as described (Okba et al. 2020). Samples were tested at a 1:101 dilution and results were considered positive above an optical density (OD) ratio of 1.1. In addition to the S1-IgA/IgG ELISA we applied a microarray-based immunoassay (SeraSpot®Anti-SARS-CoV-2 IgG, Seramun Diagnostica GmbH, Heidesee, Germany), that uses four recombinant SARS-CoV-2 proteins (complete Spike, Spike S1 domain, Spike receptor binding domain (RBD), and nucleocapsid protein), printed in an array arrangement on the bottom of each well. Antibodies were detected by use of a horseradish peroxidase-(HRP)-labeled antibody against human IgG. Color intensity at the site of formed immune complexes correlates to the antibody concentration. Results are shown as signal-to-cut-off (S/CO) ratios, obtained by dividing the signal of a specific spot by that of an internal control spot in each well. Samples showing at least two out of the four antigens S/CO ratio of ≥ 1.0 were regarded as positive (Schwarz et al. 2021).

**Viral load quantification**

SARS-CoV-2 RNA quantification was performed by real-time RT-PCR on swabs from the upper respiratory tract (URT) obtained within standard care. RNA concentrations are given as the logarithm base 10. RNA concentration was calculated from the cycle threshold (Ct) value of a RT-PCR targeting the E gene (Corman et al. 2020) and by using an empirical formula derived from testing standard curves of SARS-CoV-2 RNA and cell culture supernatants as described elsewhere (Jones et al. 2021).

**CD169/Siglec-1 expression on monocytes**

CD169/Siglec-1 expression on monocytes was analyzed as described (Stuckrad et al. 2020). Briefly, CD169/Siglec-1 expression on monocytes was analyzed in EDTA whole blood. Whenever possible, analyses were performed on the same day, within 4 h after blood samples were taken. Otherwise, EDTA blood was stored at 2–8 °C and immediately processed the next morning. Analysis was performed using an accredited flow cytometry protocol at the clinical diagnostics laboratory (Labor Berlin GmbH). Samples were incubated with a mouse anti-human antibody mixture consisting of antibodies against CD169/Siglec-1 (clone 7–239, labelled with a PE-fluorochrome/protein ratio of 1:1), CD14 (APC) and CD45 (Krome Orange, all antibodies were obtained from Beckman Coulter). Stained samples were acquired on a 10-color flow cytometer (Navios EX, Beckman Coulter) and analyzed using the Navios software. During each analytical run, QuantiBRITE™ PE tubes (BD Biosciences) were used to convert the fluorescent channel 2 (FL2) mean fluorescent intensity (MFI) signals on CD14+ monocytes to monoclonal antibodies bound per cell (mAb/cell) values. FL2 MFI values and absolute values of PE molecules (as given by the manufacturer) for each QuantiBRITE bead population were used to perform linear least square regression analysis to determine the best calibration value which then was used to convert the FL2 MFI values of monocytes in the analytical sample into the amount of PE-labelled CD169 mAb bound per monocyte (mAb/cell). The reference range for the expression of Siglec-1 in healthy controls was determined to be less than 2 500 Siglec-1 molecules/monocyte.

**Statistical analysis**

Statistical analysis was conducted with GraphPad Prism (version 9.1.2) and JMP (version 15.2.1). A p-value <0.05 was considered to be statistically significant. Continuous variables were expressed as median and interquartile ranges (IQR) and differences between groups were examined by Welch's *t* test or, in the absence of normal distribution, by Mann–Whitney *U* test. Categorical variables were compared using Chi-square tests. Kaplan-Meier curves were created using censored data and statistical testing was performed using log-rank test.

### Cohort A batch 1, IFN-α

a

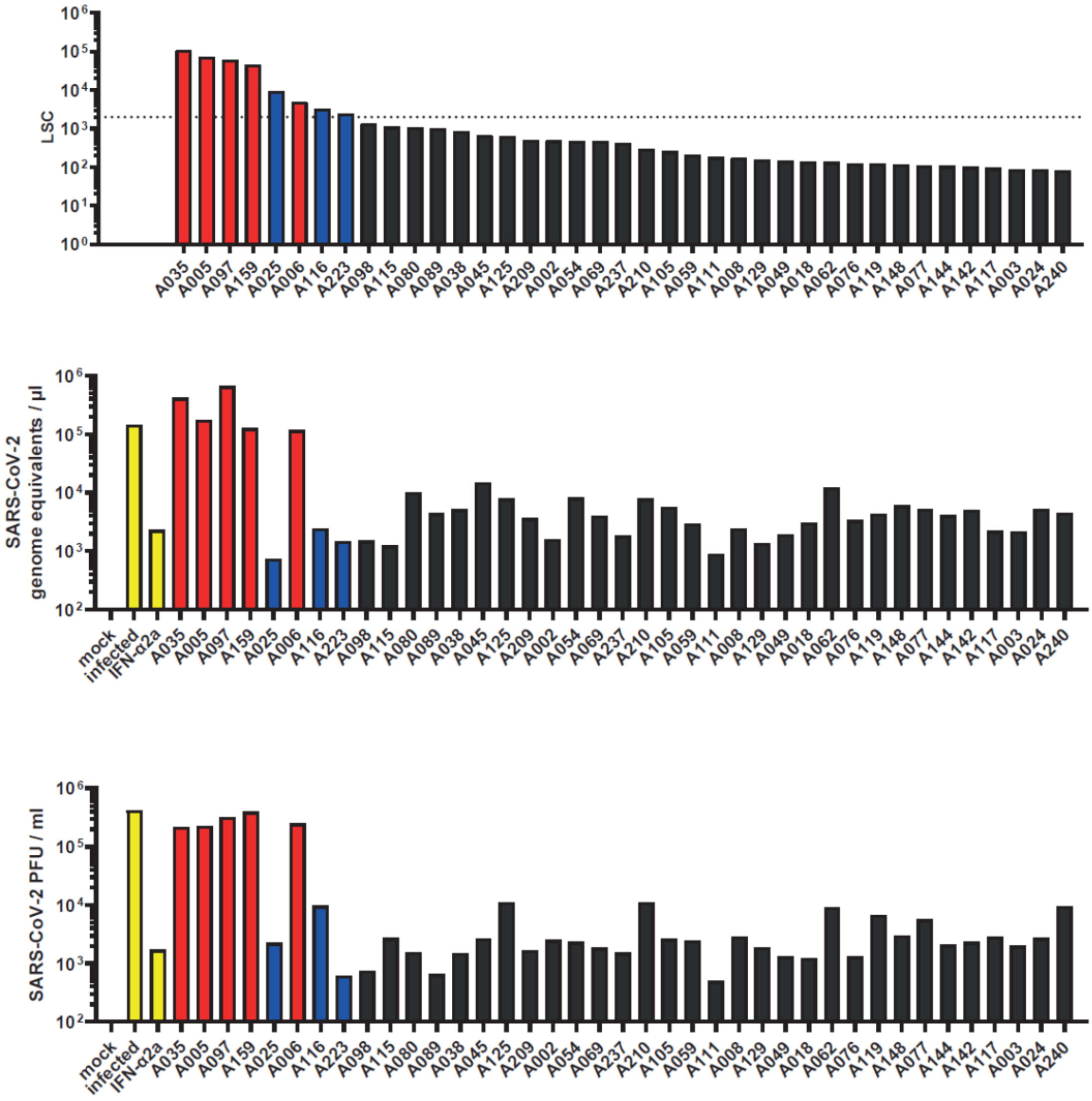

**Cohort A batch 1, IFN-ω**

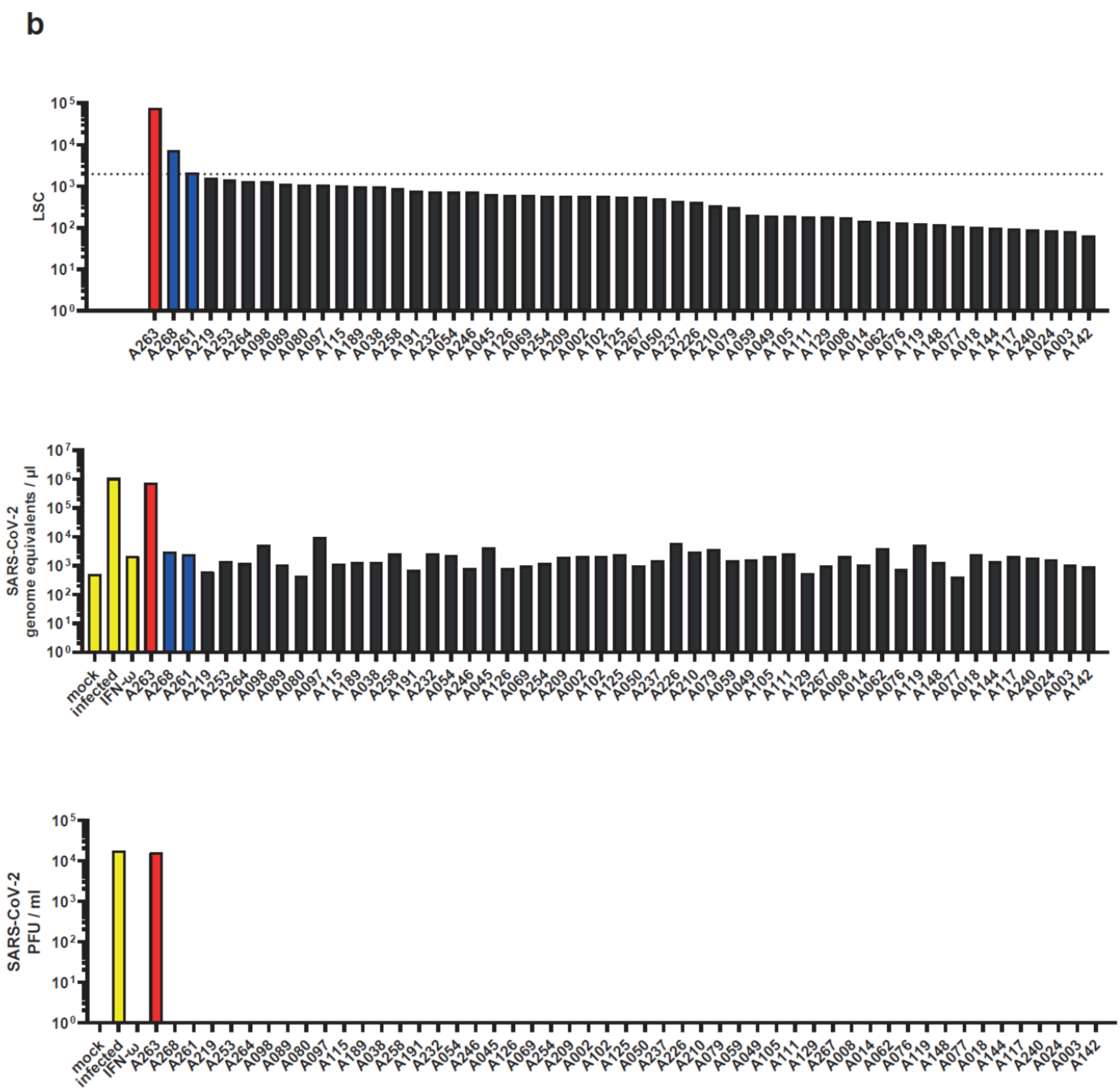

### Cohort A batch 2, IFN-α

**c**

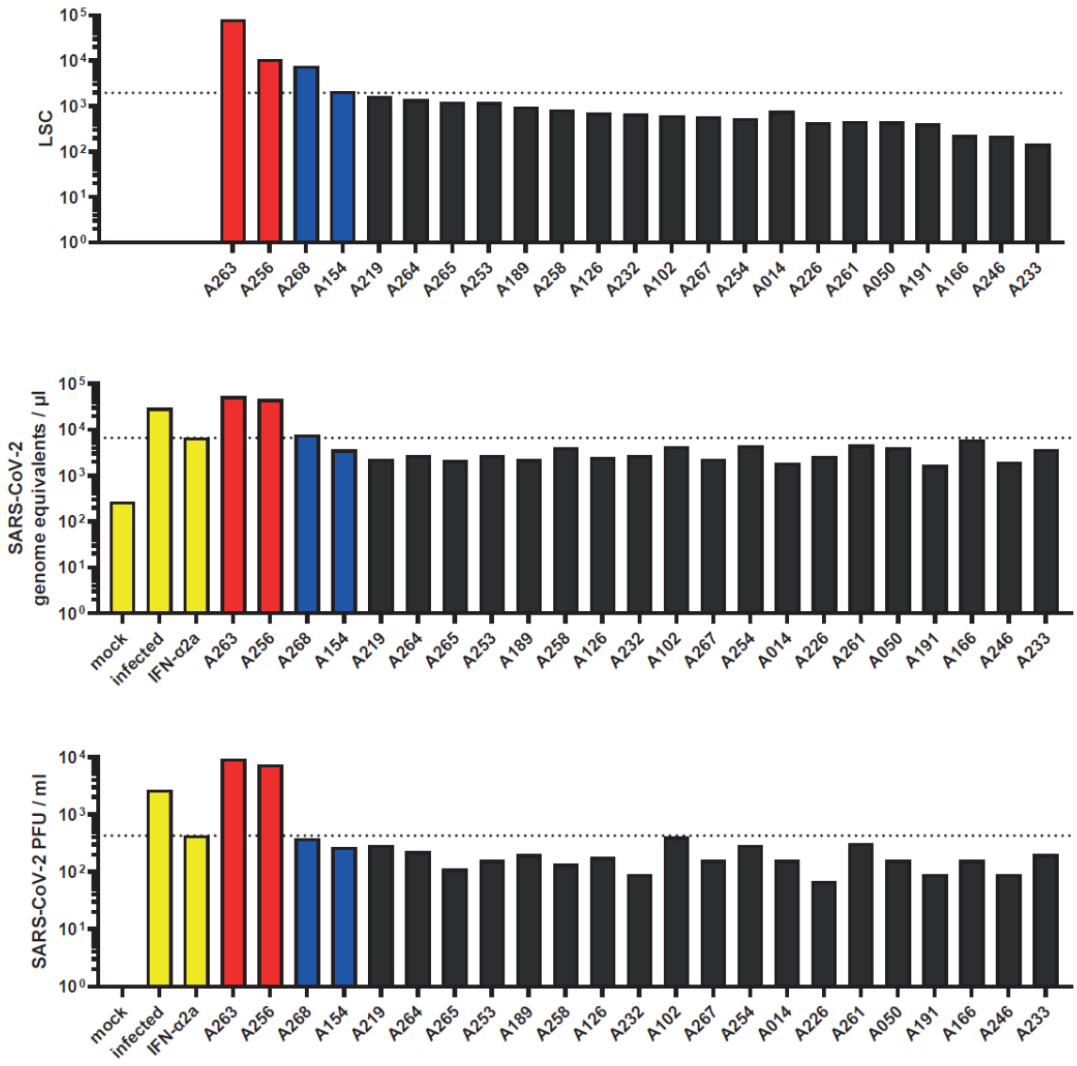

### Cohort A batch 2, IFN-ω

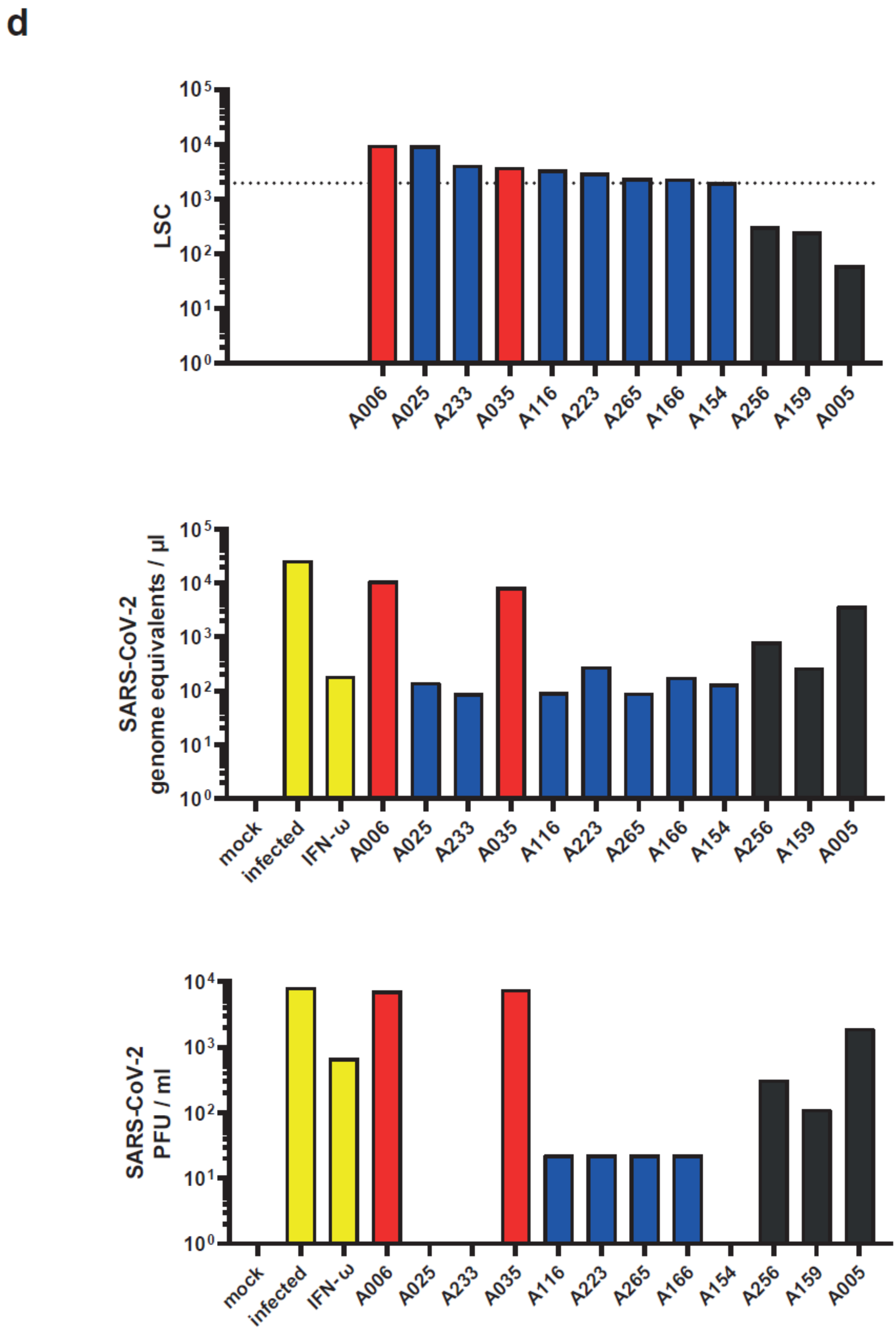

### Cohorts B and C, IFN-α

**e**

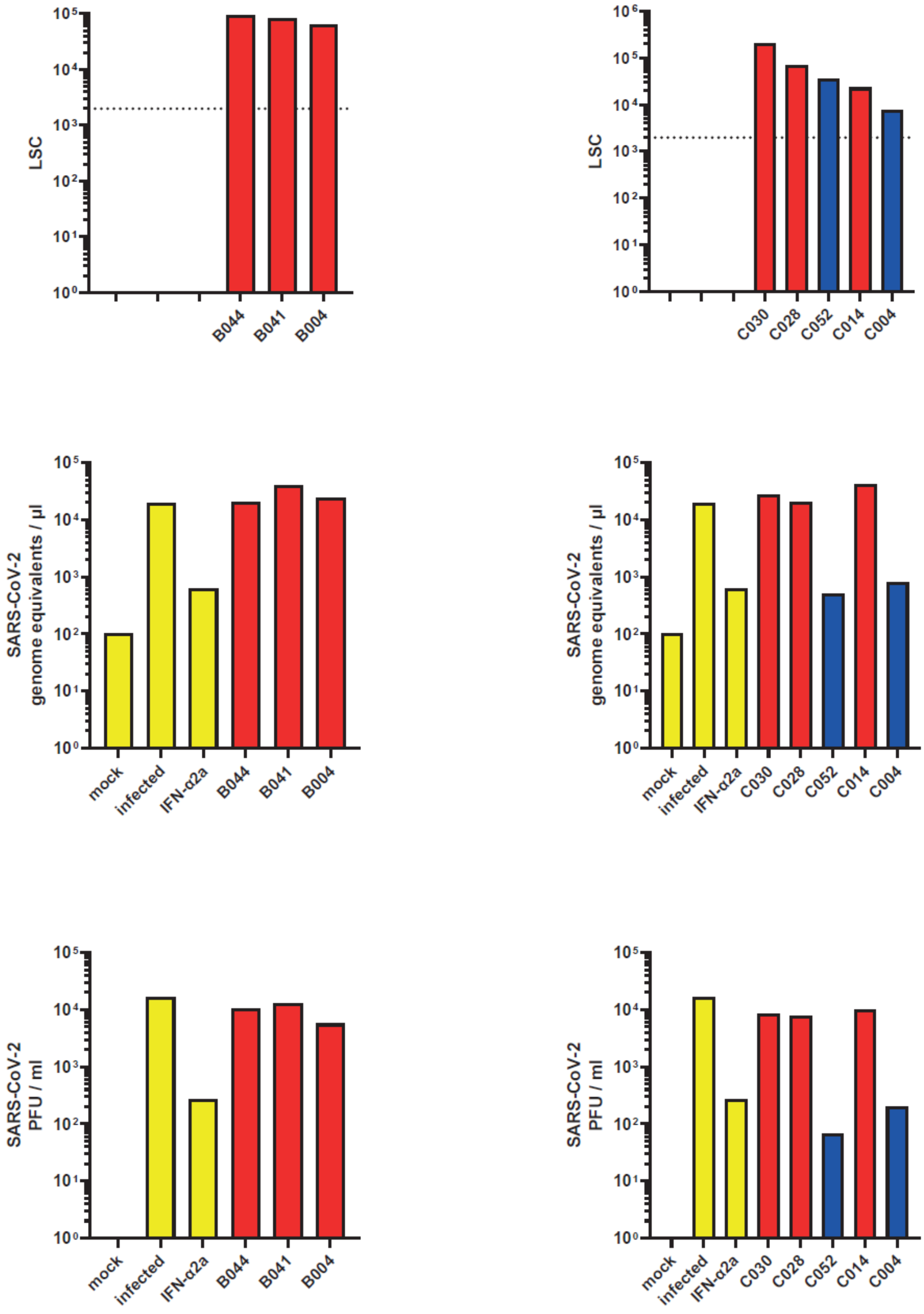

### Cohorts B and C, IFN-ω

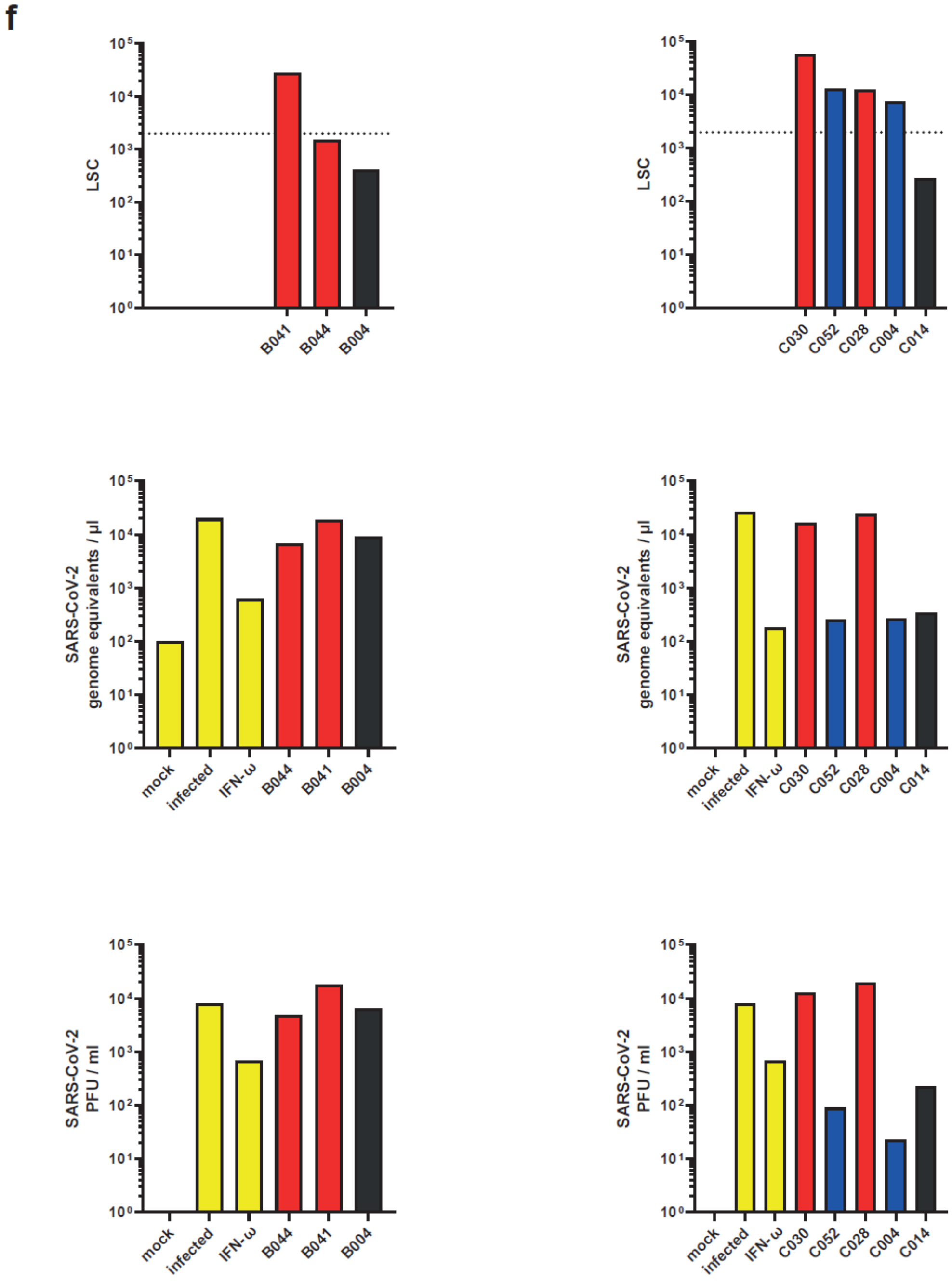

### Cohort D, IFN-α

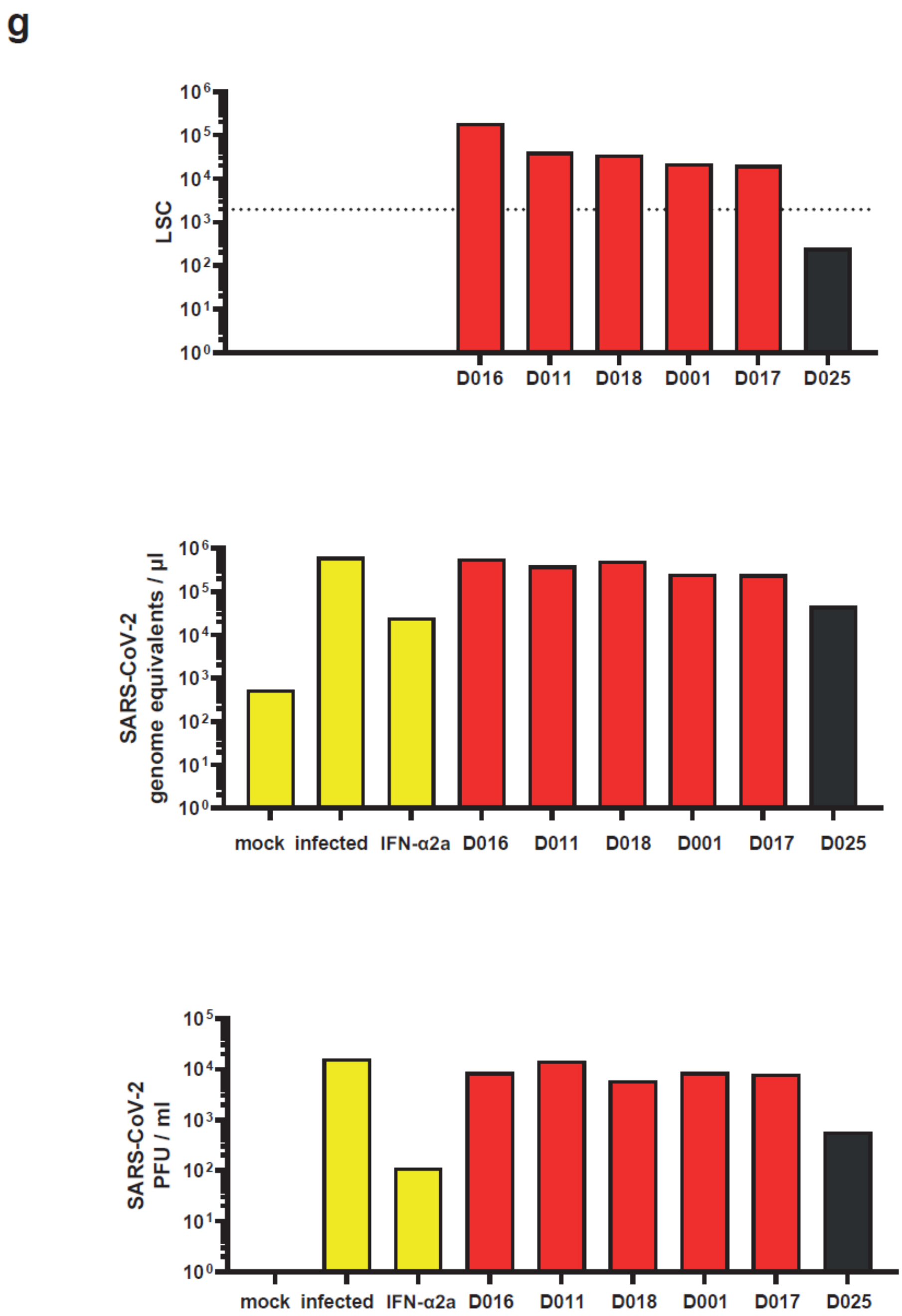

### Cohort D, IFN-ω

h

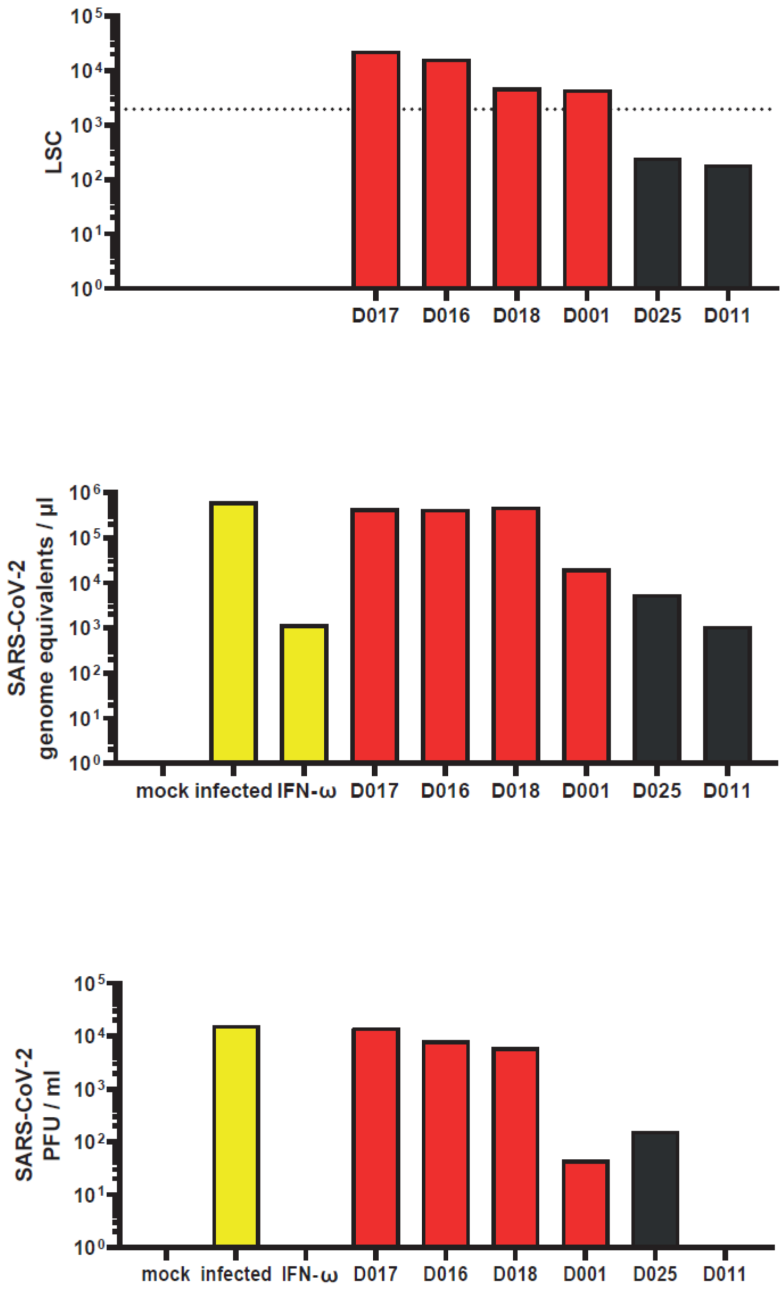

### HC, IFN-α

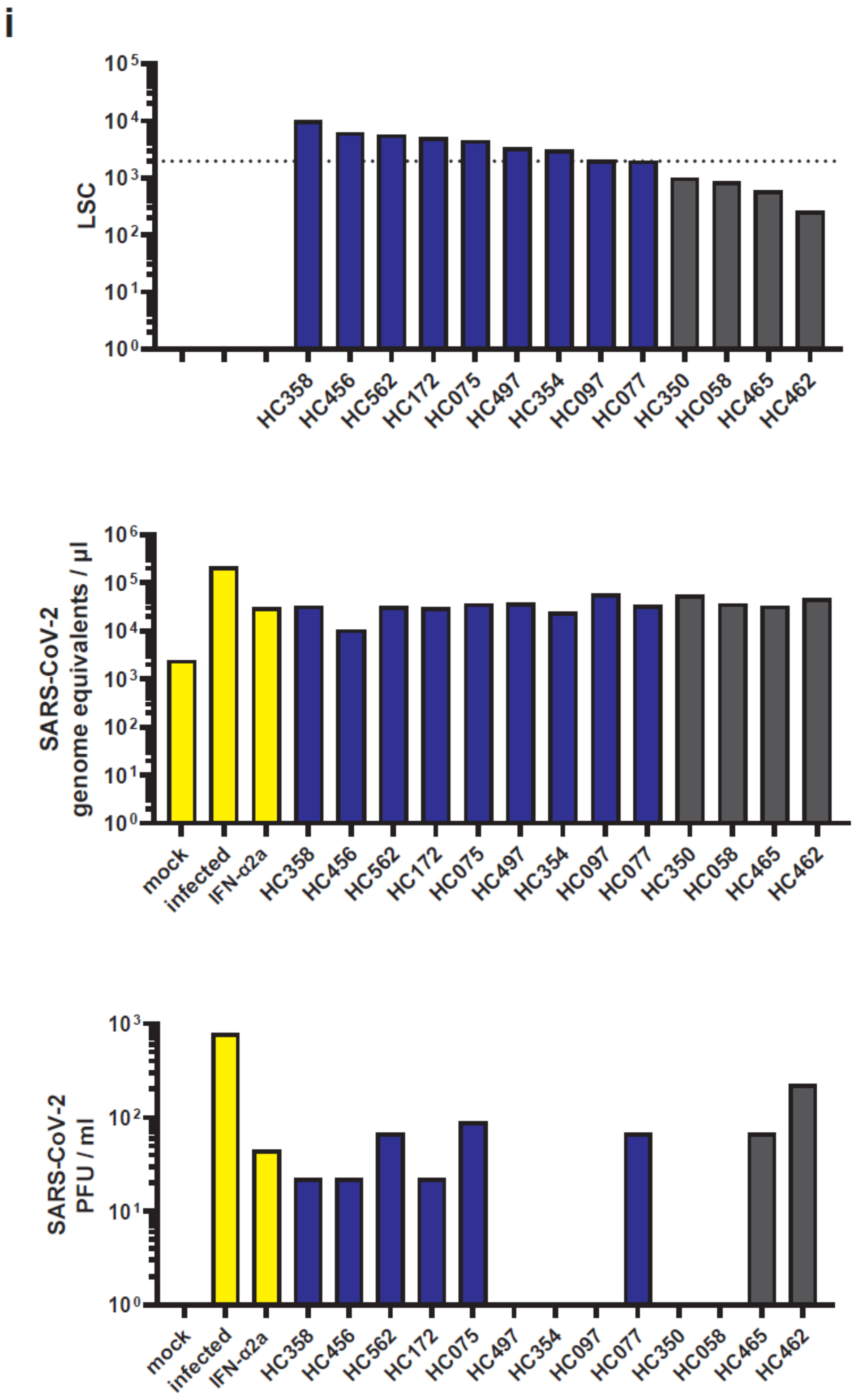
i
LSC
10⁵
10⁴
10³
10²
10¹
10⁰
HC358
HC456
HC562
HC172
HC075
HC497
HC354
HC097
HC077
HC350
HC058
HC465
HC462
SARS-CoV-2
genome equivalents / µl
10⁶
mock
infected
IFN-α2a
SARS-CoV-2
PFU / ml

### HC, IFN-ω

j

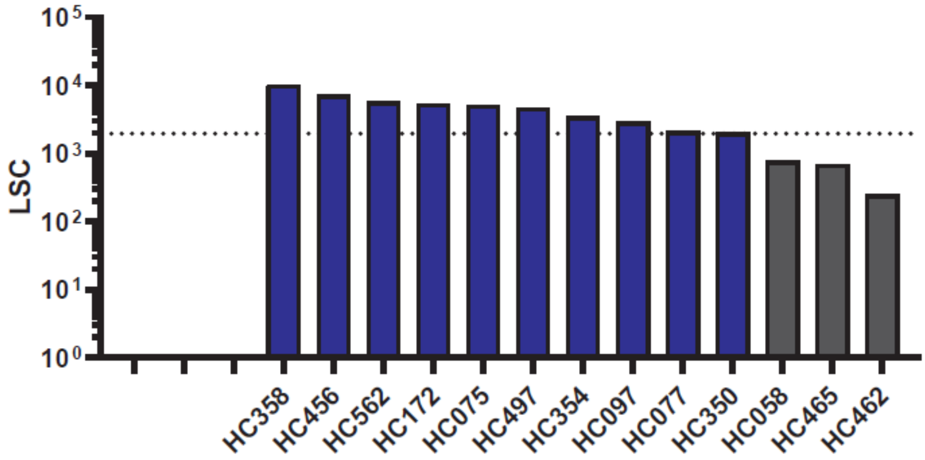
LSC
10⁰
10¹
10²
10³
10⁴
10⁵
HC358
HC456
HC562
HC172
HC075
HC497
HC354
HC097
HC077
HC350
HC058
HC465
HC462

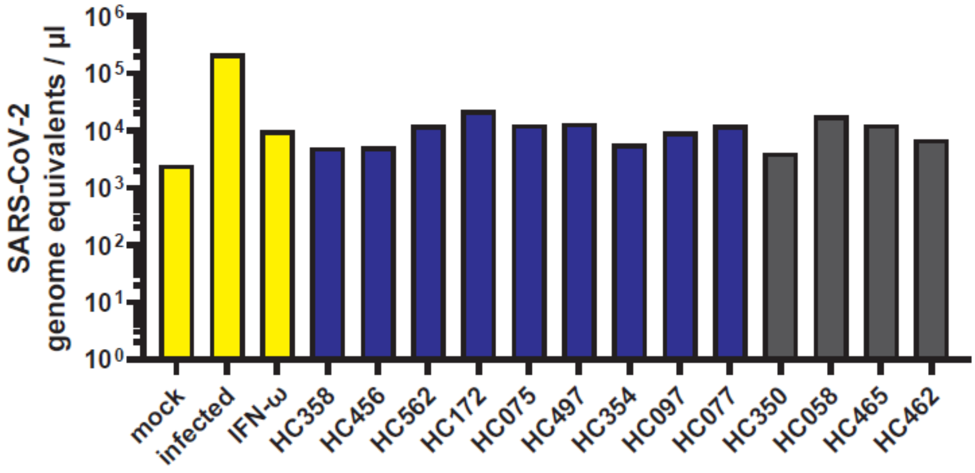
SARS-CoV-2
genome equivalents / µl
10⁰
10¹
10²
10³
10⁴
10⁵
10⁶
mock
infected
IFN-ω
HC358
HC456
HC562
HC172
HC075
HC497
HC354
HC097
HC077
HC350
HC058
HC465
HC462

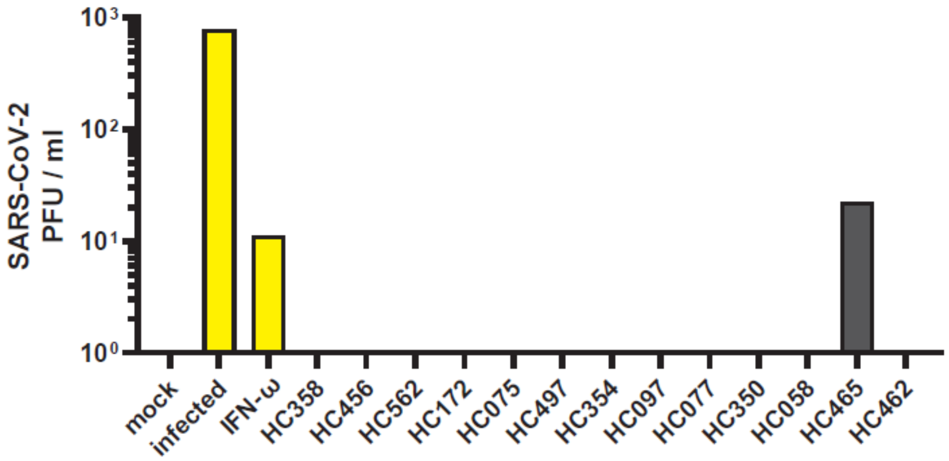
SARS-CoV-2
PFU / ml
10⁰
10¹
10²
10³
mock
infected
IFN-ω
HC358
HC456
HC562
HC172
HC075
HC497
HC354
HC097
HC077
HC350
HC058
HC465
HC462

**Figure S1. Individual test results of all tested sera**

Shown are the ECLIA assay LSC of individual sera (upper panels). Dotted line indicates the 97.5th percentile of the ECLIA assay LSC in sera from the HC (see Fig. 1).

The ability of the sera to neutralize exogenous IFN-ω is shown by the rescue of susceptibility to infection as judged by quantification of viral RNA (middle panels) and infectivity (bottom panels) in the supernatant. The infection condition in the absence of serum and IFN is set to 1.

Colored bars indicate samples containing AABs scoring specific (red) or unspecific (blue) for IFN-α2 and IFN-ω binding in the competition assay, respectively.

(**a**) Cohort A, batch 1, IFN-α

(**b**) Cohort A, batch 1, IFN-ω

(**c**) Cohort A, batch 2, IFN-α

(**d**) Cohort A, batch 2, IFN-ω

(**e**) Cohort B and C, IFN-α

(**f**) Cohort B and C, IFN-ω

(**g**) Cohort D, IFN-α

(**h**) Cohort D, IFN-ω

(**i**) HC, IFN-α

(**j**) HC, IFN-ω

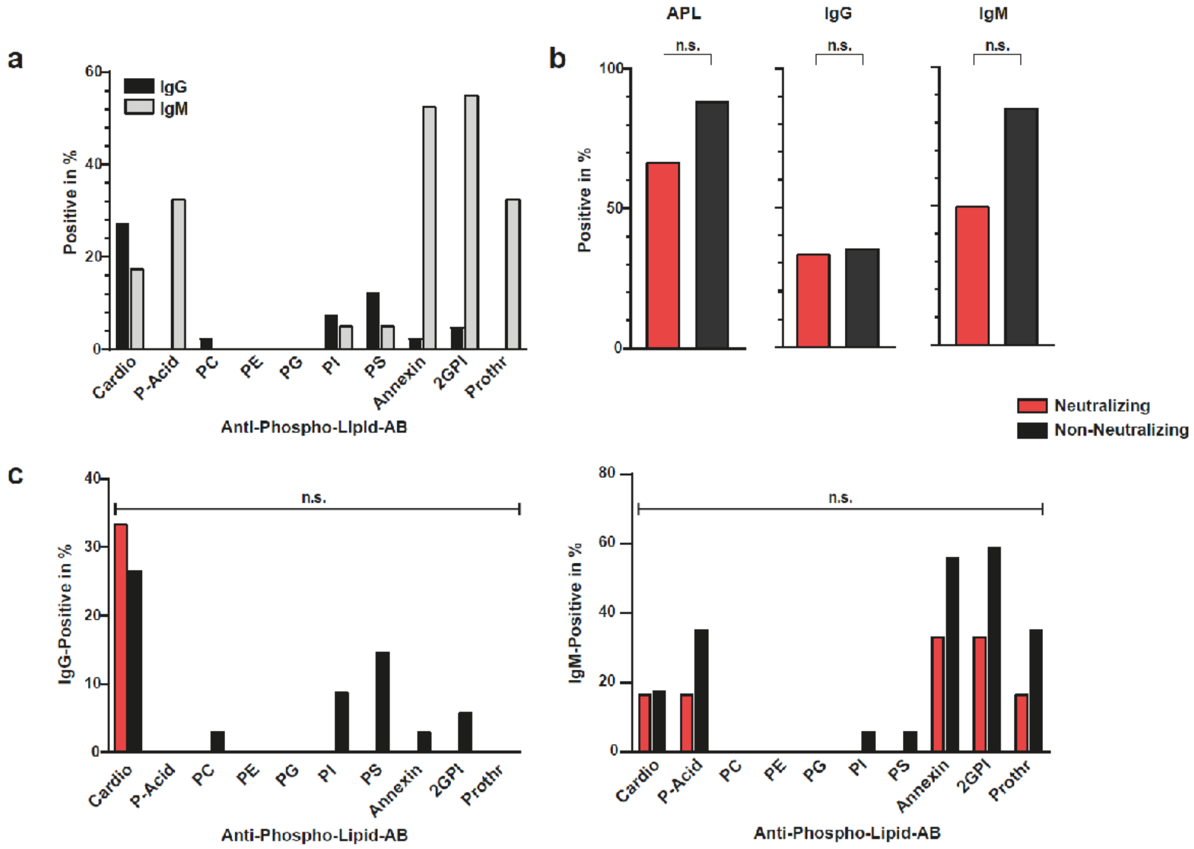


**Figure S2. Anti-phospho-lipid-antibodies (APL-ABs) in patients with and without neutralizing IFN-AABs.**

The presence of IgG and IgM antibodies to ten phospholipids was detected in a subset of patients with and without neutralizing IFN-AABs from cohort A at the peak of the disease using an immunoassay. All available samples from patients with neutralizing IFN-AAB (N=6, red) and without neutralizing IFN-AAB (N=34) were analyzed.

(**a**) Proportion of APL-AB (IgG and IgM) from all patients.

(**b**) Proportion of sera with any APL-AB (IgG and/or IgM) detected in the two groups.

(**c**) Proportion of patients with APL-ABs of the IgG and IgM isotype against the ten different phospholipids in the two groups.

Statistical testing was performed using the Chi-Square test.

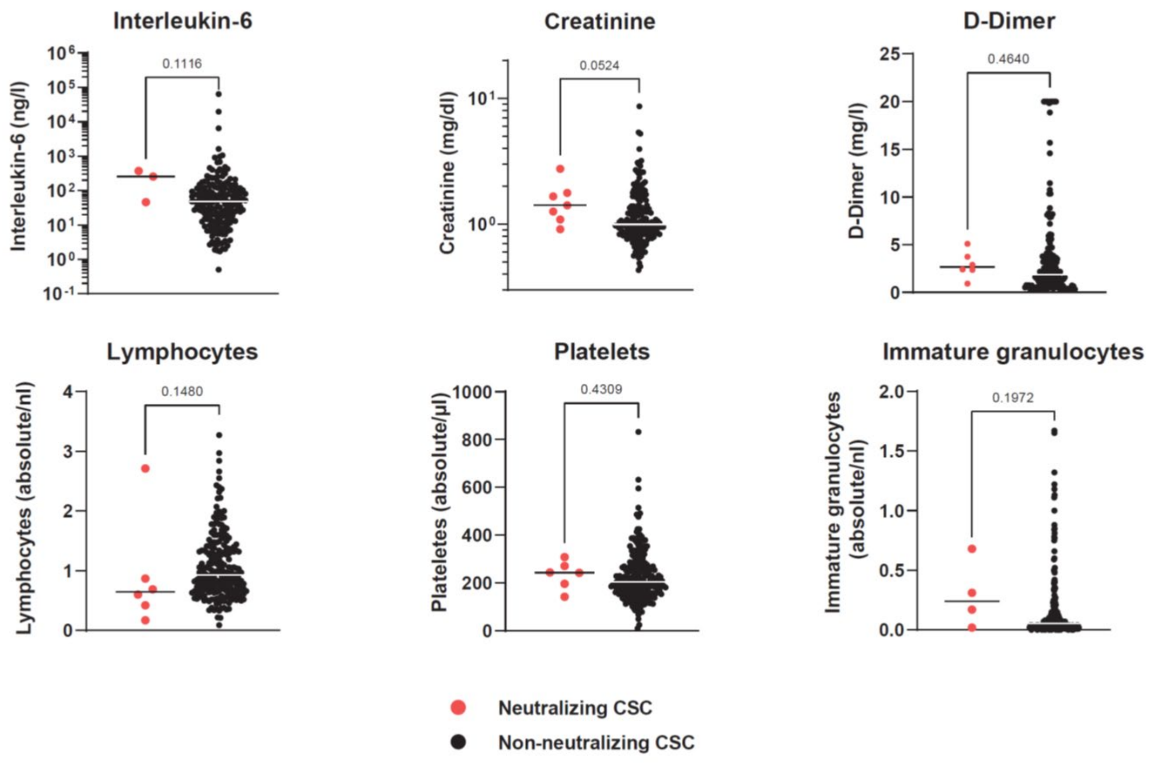


**Figure S3. Additional laboratory parameters from the CSC.** Data are shown in two groups (neutralizing N= 3-7; non-neutralizing N = 170-267). For each patient, the first available parameter within 72 hours of hospital admission is shown. Statistical testing was performed using Mann-Whitney-U test.

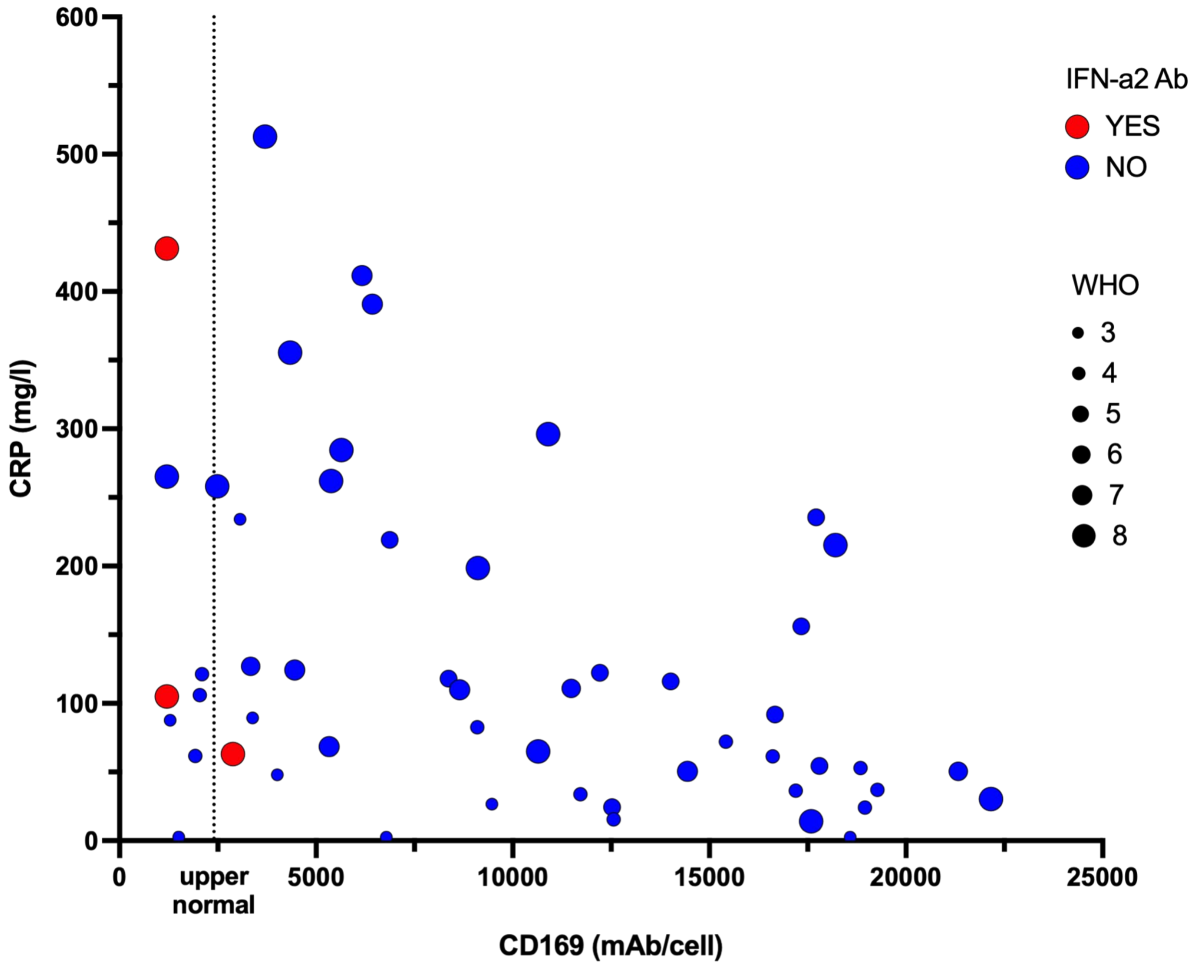


**Figure S4. Low levels of type I IFN-induced CD169/Siglec-1 on monocytes in patients with neutralizing IFN-AABs**

The expression of the type I IFN-induced protein CD169/Siglec-1 on monocytes was analyzed by flow cytometry within the first three days after hospital admission in a subgroup of patients of cohort A and plotted against corresponding CRP levels. Patients with neutralizing IFN-a2 AABs are shown by red dots, while blue dots represent patients without anti-IFN AABs. Disease severity (WHO score) is indicated by increasing dot size. The upper normal value for CD169/Siglec-1 expression on monocytes (<2400 mAb/cell) is shown by the dotted line.

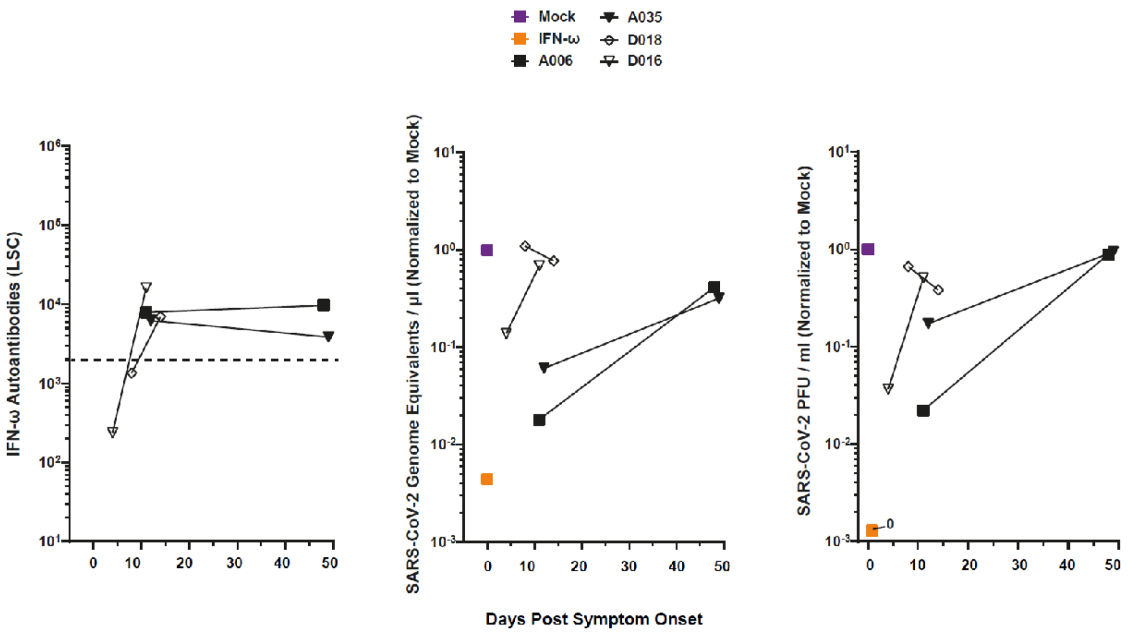


**Figure S5. In IFN-AAB-positive patients, days early PSO and at the peak of the disease share presence of high quantities of neutralizing IFN-omega AAB**

(**a**) Time course of antibody quantities in patient sera that scored IFN-AAB positive at the peak of disease (N=4). The dotted line indicates the 97.5th percentile of the ECLIA assay LSC in sera from the HC (see Fig. 1).

The ability of the sera to neutralize exogenous IFN-□ is shown by the rescue of susceptibility to infection as judged by quantification of viral RNA (**b**) and infectivity (**c**) in the supernatant. The infection condition in the absence of serum and IFN is set to 1.

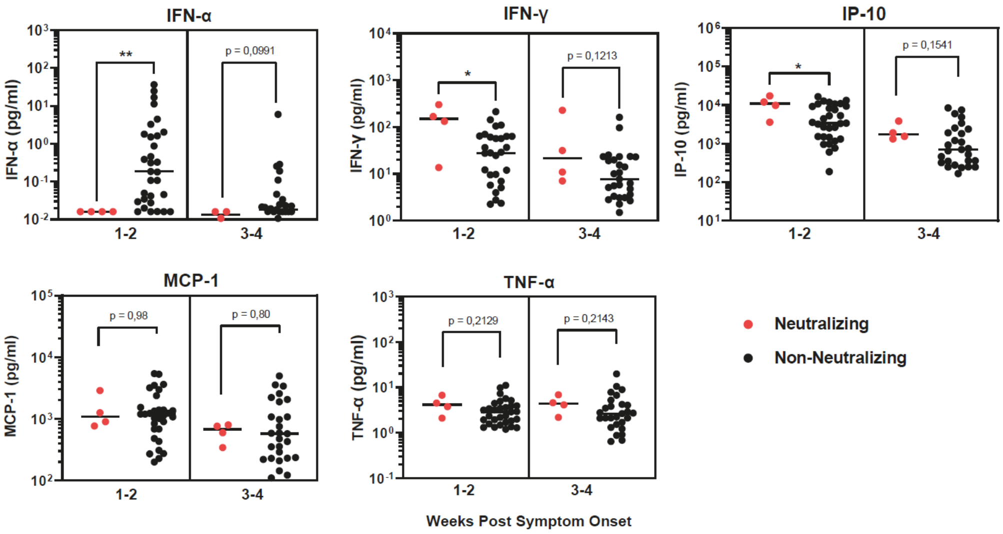


**Figure S6. Cytokine levels in sera from patients with IFN-AABs**

Sera from a subset of patients with critical COVID-19 (WHO max. 6-8) within the first four weeks PSO from cohort A were analyzed. Samples from patients with (N=4) and without (N=30-31) neutralizing IFN-AAB were compared. When more than one sample per time point (week 1-2 or 3-4) was available, the mean from all samples was calculated and plotted. Statistical testing was performed using the Mann-Whitney *U* test.

**a**

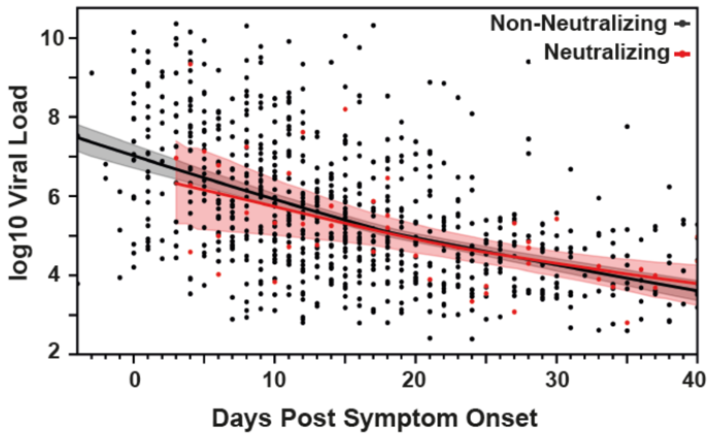
Non-Neutralizing
Neutralizing
log10 Viral Load
10
8
6
4
2
0
10
20
30
40
Days Post Symptom Onset

**b**

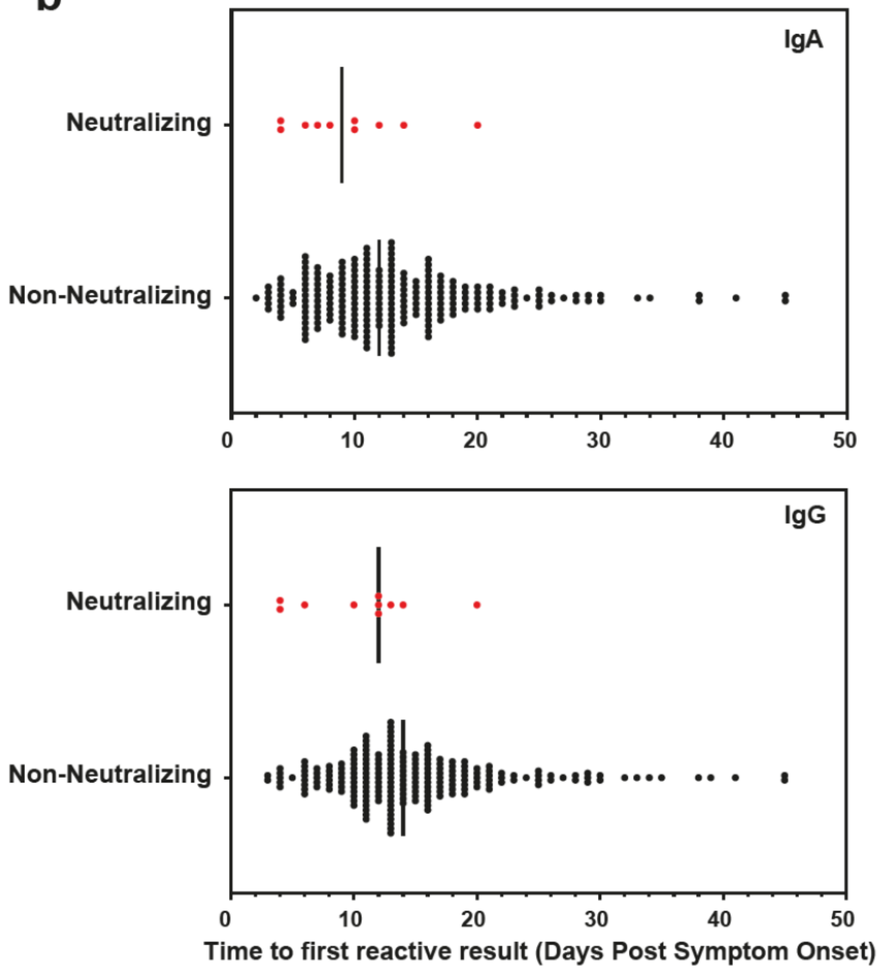
IgA
Neutralizing
Non-Neutralizing
0
10
20
30
40
50
IgG
Neutralizing
Non-Neutralizing
0
10
20
30
40
50
Time to first reactive result (Days Post Symptom Onset)

**Figure S7. SARS-CoV-2 RNA load and time to first reactive IgA or IgG result**

(**a**) Viral RNA concentration over time in upper respiratory tract (URT) samples in two groups of hospitalized COVID-19 patients from cohort A and D, neutralizing patients in red (N=10) and non-neutralizing patients in black (N=225 for IgA, N=205 for IgG). Bold lines and shaded areas indicate the LOWESS fit and 95% confidence intervals for the two groups, respectively. (**b**) Time to first reactive IgG or IgA ELISA result PSO.

Median time to first reactive IgA result: Neutralizing 9 days, non-neutralizing 12 days.

Median time to first reactive IgG result: Neutralizing 12 days, non-neutralizing 14 days.

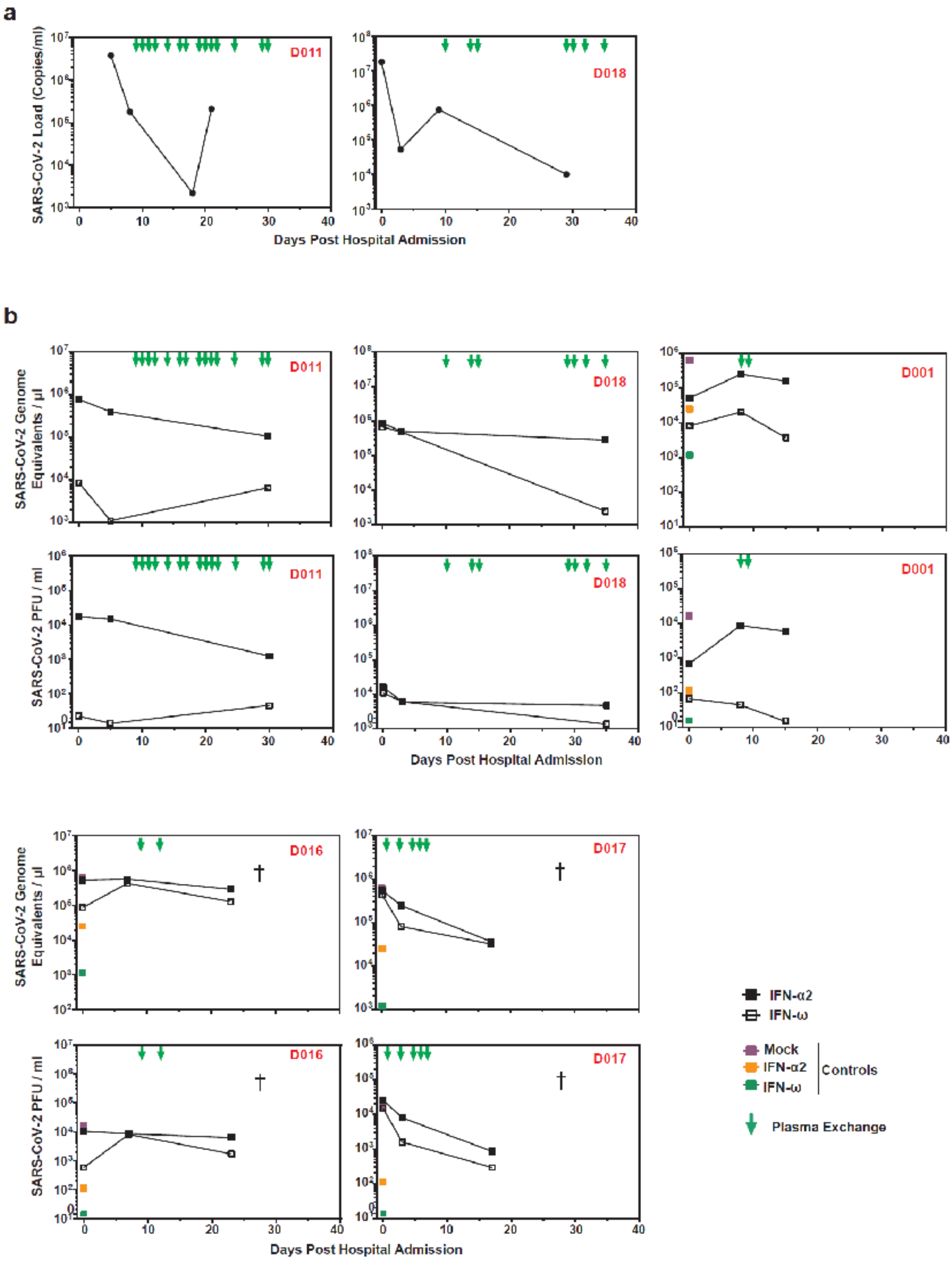


**Figure S8. Viral load (a) and neutralization data (b) of COVID-19 patients undergoing TPE**

**Supplementary Table 1. COVIMMUN Clinical Characteristics**

| | **All participants** |
|---|---|
| **Number of participants** | 667 |
| **Age** (Median, IQR[1], n/N) | 37 (30-49; 653/667) |
| **Sex**<br>Female (%, n/N)<br>Male (%, n/N) | <br>65.2 (423/649)<br>34.8 (226/649) |
| **BMI[2]** (kg/m$^2$, Median, IQR, n/N) | 23.9 (21.3-26.6; 624/667) |
| **Comorbidities**<br>Cardiovascular disease (%, n/N)<br>Pulmonary disease (%, n/N)<br>Diabetes (%, n/N)<br>Obesity (%, n/N)<br>Autoimmune disease (%) | <br>10.4 (68/651)<br>6.1 (40/651)<br>1.4 (9/651)<br>12.2 (76/624)<br>3.4 (22/651) |

[1]Interquartile range

[2]Body mass index

**Supplementary Table 2. Raw data of AAB quantification**

| | | | ELISA LSC | | Competition | | Neutralization | |
|---|---|---|---|---|---|---|---|---|
| Patient ID | Days PSO | Days post hospital admission | IFN-α | IFN-□ | IFN-α | IFN-□ | IFN-α | IFN-□ |
| A005 | 17 | 11 | 29290 | 330 | - | - | YES | NO |
| A005 | 26 | 20 | 70266 | 61 | YES | NO | YES | NO |
| A006 | 11 | 8 | 13035 | 7897 | - | - | NO | NO |
| A006 | 48 | 45 | 4740 | 9689 | YES | YES | YES | YES |
| A035 | 12 | 10 | 58196 | 6221 | - | - | YES | NO |
| A035 | 49 | 47 | 105231 | 3844 | YES | YES | YES | YES |
| A097 | 12 | 9 | 59667 | 1070 | YES | NO | YES | NO |
| A159 | 8 | 8 | 61708 | 208 | - | - | YES | NO |
| A159 | 24 | 24 | 44086 | 254 | YES | NO | YES | NO |
| A256 | 20 | 20 | 32944 | 677 | - | - | YES | NO |
| A256 | 43 | 43 | 10858 | 316 | YES | NO | YES | NO |
| A263 | 29 | 25 | 79784 | 77465 | YES | YES | YES | YES |
| B004 | 21 | 7 | 63854 | 410 | YES | NO | YES | YES |
| B041 | 12 | 3 | 81745 | 27058 | YES | YES | YES | YES |
| B044 | 19 | 6 | 94109 | 1493 | YES | YES | YES | YES |

| | | | | | | | | |
|---|---|---|---|---|---|---|---|---|
| C014 | 11 | 5 | 22819 | 264 | YES | NO | YES | NO |
| C028 | no data | 15 | 67408 | 12292 | YES | YES | YES | YES |
| C030 | 14 | 10 | 203323 | 55886 | YES | YES | YES | YES |
| D001 | 6 | 0 | 3723 | 2479 | - | - | NO | NO |
| D001 | 12 | 6 | 22589 | 4479 | YES | YES | YES | NO |
| D001 | 13 | 7 | 7670 | 2845 | - | - | - | - |
| D001 | 14 | 8 | 2954 | 1611 | - | - | - | - |
| D001 | 19 | 13 | 11238 | 9378 | - | - | YES | NO |
| D011 | 6 | 0 | 10377 | 135 | - | - | YES | NO |
| D011 | 16 | 10 | 40642 | 190 | YES | NO | YES | NO |
| D011 | 17 | 11 | 15888 | 205 | - | - | - | - |
| D011 | 18 | 12 | 9481 | 217 | - | - | - | - |
| D011 | 19 | 13 | 5642 | 258 | - | - | - | - |
| D011 | 20 | 14 | 2367 | 260 | - | - | - | - |
| D011 | 21 | 15 | 3279 | 269 | - | - | - | - |
| D011 | 23 | 17 | 2050 | 196 | - | - | - | - |
| D011 | 27 | 21 | 389 | 206 | - | - | - | - |
| D011 | 30 | 24 | 749 | 327 | - | - | - | - |
| D011 | 35 | 29 | 611 | 219 | - | - | - | - |

| | | | | | | | | |
|---|---|---|---|---|---|---|---|---|
| D011 | 36 | 30 | 589 | 265 | - | - | NO | NO |
| D016 | 4 | 0 | 28107 | 237 | - | - | YES | NO |
| D016 | 11 | 7 | 190185 | 16167 | YES | YES | YES | YES |
| D016 | 13 | 9 | 108701 | 6140 | - | - | - | - |
| D016 | 16 | 12 | 134202 | 9771 | - | - | - | - |
| D016 | 17 | 13 | 79312 | 3932 | - | - | - | - |
| D016 | 23 | 19 | 89935 | 6232 | - | - | - | - |
| D016 | 27 | 23 | 90694 | 5413 | - | - | YES | NO |
| D017 | 10 | 0 | 11136 | 22968 | - | - | YES | YES |
| D017 | 13 | 3 | 20609 | 39264 | YES | YES | YES | NO |
| D017 | 14 | 4 | 5569 | 7678 | - | - | - | - |
| D017 | 15 | 5 | 1649 | 2462 | - | - | - | - |
| D017 | 16 | 6 | 1075 | 1079 | - | - | - | - |
| D017 | 17 | 7 | 540 | 598 | - | - | - | - |
| D017 | 20 | 10 | 1189 | 1445 | - | - | - | - |
| D017 | 27 | 17 | 1487 | 1948 | - | - | NO | NO |
| D018 | 8 | 0 | 34379 | 6957 | YES | YES | YES | YES |
| D018 | 14 | 6 | 35790 | 4804 | - | - | YES | YES |
| D018 | 18 | 10 | 7222 | 350 | - | - | - | - |

| | | | | | | | | |
|---|---|---|---|---|---|---|---|---|
| D018 | 21 | 13 | 11518 | 596 | - | - | - | - |
| D018 | 22 | 14 | 4075 | 300 | - | - | - | - |
| D018 | 23 | 15 | 2240 | 361 | - | - | - | - |
| D018 | 35 | 27 | 6480 | 289 | - | - | - | - |
| D018 | 37 | 29 | 2077 | 325 | - | - | - | - |
| D018 | 38 | 30 | 742 | 377 | - | - | - | - |
| D018 | 40 | 32 | 517 | 160 | - | - | - | - |
| D018 | 42 | 34 | 1368 | 152 | - | - | - | - |
| D018 | 43 | 35 | 371 | 194 | - | - | YES | NO |
